# Transdiagnostic failure to adapt interoceptive precision estimates across affective, substance use, and eating disorders: A replication study

**DOI:** 10.1101/2023.10.11.23296870

**Authors:** Claire A. Lavalley, Navid Hakimi, Samuel Taylor, Rayus Kuplicki, Katherine L. Forthman, Jennifer L. Stewart, Martin P. Paulus, Sahib S. Khalsa, Ryan Smith

## Abstract

Recent computational theories of interoception suggest that perception of bodily states rests upon an expected reliability- or precision-weighted integration of afferent signals and prior beliefs. The computational psychiatry framework further suggests that aberrant precision-weighting may lead to misestimation of bodily states, potentially hindering effective visceral regulation and promoting psychopathology. In a previous study, we fit a Bayesian computational model of perception to behavior on a heartbeat tapping task to test whether aberrant precision-weighting was associated with misestimation of bodily states. We found that, during an interoceptive perturbation designed to amplify afferent signal precision (inspiratory breath-holding), healthy individuals increased the precision-weighting assigned to ascending cardiac signals (relative to resting conditions), while individuals with symptoms of anxiety, depression, substance use disorders, and/or eating disorders did not. A second study also replicated the pattern observed in healthy participants. In this pre-registered study, we aimed to replicate our prior findings in a new transdiagnostic patient sample (N=285) similar to the one in the original study. These new results successfully replicated those found in our previous study, indicating that, transdiagnostically, patients were unable to adjust beliefs about the reliability of interoceptive signals – preventing the ability to accurately perceive changes in their bodily state. Follow-up analyses combining samples from the previous and current study (N=719) also afforded the power to identify group differences within narrower diagnostic groups and to examine predictive accuracy when logistic regression models were trained on one sample and tested on the other. Given the increased confidence in the generalizability of these effects, future studies should examine the utility of interceptive precision measures in predicting treatment outcomes or identify whether these computational mechanisms might represent novel therapeutic targets for improving visceral regulation.

## Introduction

Interoception – the process by which the nervous system senses, interprets, and integrates the internal state of the body – is thought to play an important role in a number of psychiatric disorders. Evidence of altered interoceptive processing has been observed in depression, anxiety, eating, and substance use disorders, among others (reviewed in (1)). This incudes, for example, negative associations between depression and heartbeat counting accuracy (2–5), heightened interoceptive sensations in panic disorder (reviewed in (6)), blunted neural responses during interoceptive processing in substance users (7), and stronger effects of expectation on interoception in eating disorders (8). However, debates about the limitations of current interoception measures have grown in recent years (9–14), and the pathophysiological mechanisms underlying psychiatric disorders remain unclear.

In an effort to address these methodological and inferential limitations, we previously applied a computational modeling approach designed to minimize certain limitations of previous approaches and to allow inferences regarding the algorithmic processing mechanisms spanning depression, anxiety, eating, and substance use disorders (15). Our model assumed that the brain updates beliefs about the state of the body by integrating afferent interoceptive signals with prior beliefs in a manner weighted by their expected reliability – or precision – corresponding to approximate Bayesian inference. As the reliability of prior beliefs and afferent signals can change over time and context, adaptive interoceptive processing requires continual tuning of these precision weightings – a process that could be affected in psychopathology. Fitting our model to behavioral data in a cardiac interoception (heartbeat tapping) task revealed that healthy individuals successfully adapted the precision assigned to both prior expectations and afferent signals during different task contexts. Most importantly in the present context, they assigned low precision to the sensory signal at rest, while this precision assignment increased during a breath-hold perturbation designed to amplify the strength of the afferent signal. In a follow-up study with a new sample of healthy individuals, we replicated this effect using the same task and model (16). This increase in precision assignment during an interoceptive perturbation was not unexpected, as only roughly 35% of individuals appear to accurately perceive their own heartbeats at rest (6). In contrast, when visceral states are perturbed, cardiac perception becomes more accurate, particularly under conditions of heightened cardiorespiratory arousal (17–19).

In contrast to healthy participants, the original study found evidence that a transdiagnostic psychiatric sample did not update their signal precision estimates during the breath-hold manipulation, suggesting rigidity in these estimates and an insensitivity to changes in the afferent signal. However, this finding remained to be replicated. In the present pre-registered study (https://osf.io/9znsg), we attempted to replicate this inflexible pattern of precision estimation in a new transdiagnostic sample with a similar composition to the previous study and who performed the same task under identical conditions.

## Methods

### Participants

Data were collected as part of the larger Tulsa 1000 project, which includes an exploratory and confirmatory dataset comprised of 1050 individuals (20). Our prior study used the exploratory dataset (N = 434), while this study used the confirmatory dataset. The available subset of the confirmatory sample (N = 479) in the Tulsa 1000 study included 94 healthy comparisons (HCs; 37 male; mean age: 32.35, SD = 11.16), 10 individuals with anxiety disorders alone (iANX; 1 male; mean age: 37.18, SD = 6.37), 57 with major depression alone (iDEP; 20 male; mean age: 36.00, SD = 10.12), 127 with co-morbid anxiety/depression (iDEP+ANX; 26 male; mean age: 31.99, SD = 9.83), 155 with substance use disorders (iSUDs; i.e., with or without co-morbid affective disorders; 60 male; mean age: 34.07, SD = 8.45), and 36 with eating disorders (iEDs; i.e., with or without co-morbid affective disorders; 3 male; mean age: 27.75, SD = 8.46). Please note that acronyms here include “i” to indicate “individuals with” so as not to identify individuals by their diagnoses in subsequent use.

To arrive at this sample, individuals aged 18-55 years were screened based on dimensional psychopathology scores. Inclusion used the following measures and criteria: Patient Health Questionnaire (PHQ-9; (21)) ≥ 10, Overall Anxiety Severity and Impairment Scale (OASIS; (22)) ≥ 8, Drug Abuse Screening Test (DAST-10; (23)) score > 2, and/or Eating Disorder Screen (SCOFF; (24)) score ≥ 2 (for screening measure scores on the full sample, see **Table 1**). Participants were grouped based on DSM-IV or DSM-5 diagnosis using the Mini International Neuropsychiatric Inventory 6 or 7 (MINI; (25)). HCs did not show elevated symptoms or possess any psychiatric diagnoses.

**Table 1.**
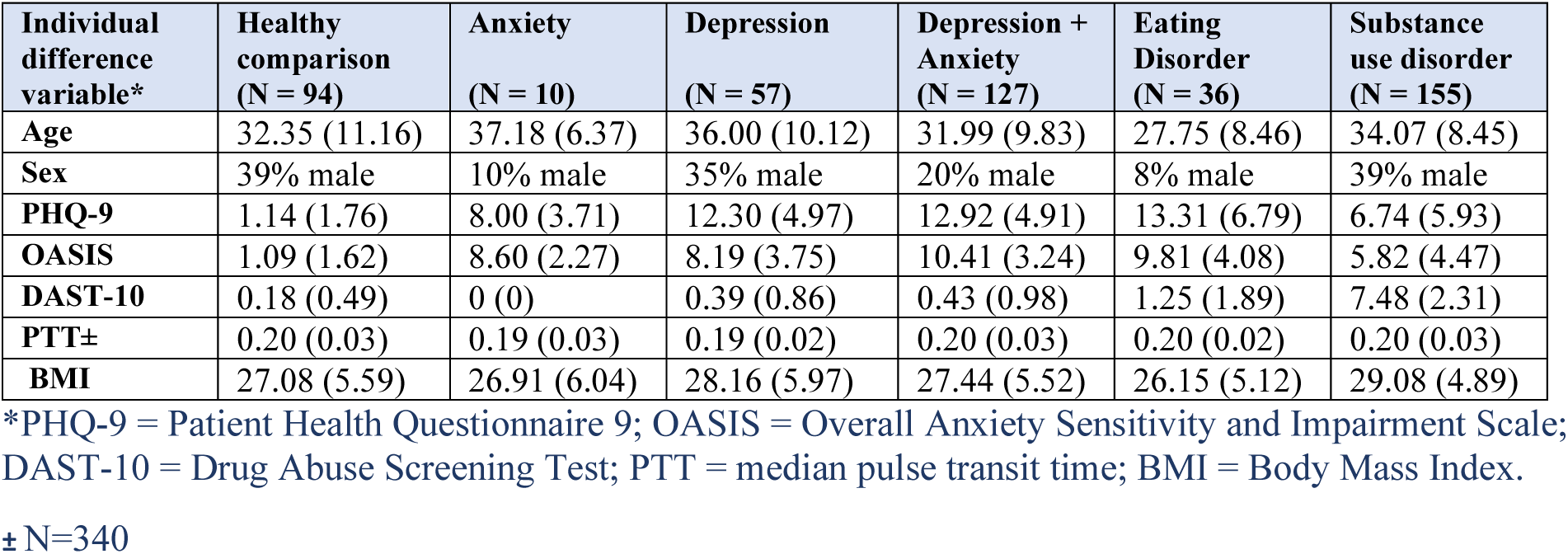
Mean (and standard deviation) for clinical and demographic variables in the full sample.

Participants were excluded if they (i) tested positive for drugs of abuse, (ii) met criteria for psychotic, bipolar, or obsessive-compulsive disorders, or reported (iii) history of moderate-to-severe traumatic brain injury, neurological disorders, or severe or unstable medical conditions, (iv) active suicidal intent or plan, or (v) change in psychotropic medication status within 6 weeks. Full inclusion/exclusion criteria are described in (20). The study was approved by the Western Institutional Review Board. All participants provided written informed consent prior to completion of the study protocol, in accordance with the Declaration of Helsinki, and were compensated for participation. ClinicalTrials.gov identifier: #NCT02450240.

The clinical and demographic information reported in **Table 1** includes the 479 participants with available data from the confirmatory dataset, excluding only those with invalid task or electrocardiogram (EKG) data. We note here, however, that some of these participants did not pass additional quality control checks and were subsequently removed, as described further below.

### Heartbeat perception task

As part of the T1000 project, participants completed several assessments, self-report measures, and behavioral tasks (detailed in (20)). The heartbeat tapping task relevant to the present study was based on a previously developed task (26, 27). As in our previous study, the task was repeated under three conditions (each for a period of 60 seconds) designed to influence prior confidence and afferent signal precision. In the “guessing” condition, participants were simply instructed to close their eyes and press down on a key when they felt their heartbeat, to try to mirror their heartbeat as closely as possible, and told that they should take their best guess even if they weren’t sure. In the “no guessing” condition, all instructions were identical except that they were told to only press the key when they actually felt their heartbeat, and if they did not feel their heartbeat that they should not press the key. This can be seen as altering prior beliefs associated with confidence thresholds (i.e., higher confidence was required to choose to press the key). In the “breath-hold” (perturbation) condition, participants were again instructed not to guess, but were also asked to first empty their lungs of all air and then inhale as deeply as possible and hold it for as long as they could tolerate (up to the length of the one-minute trial). This manipulation attempted to putatively increase the strength of the afferent cardiac signal by increasing physiological arousal. Conceptually, therefore, one can view the no-guessing instruction as a baseline condition, where adding the breath-hold increases signal precision and adding the guessing instruction instead increases prior bias toward detecting heartbeats.

An identical “tone tapping” condition was used as a control, where participants were instructed to press the key every time they heard a 1000Hz auditory tone, each of which was presented for 100ms (78 tones, randomly jittered by +/- 10% and presented in a pattern following a sine curve with a frequency of 13 cycles/minute, mimicking the range of respiratory sinus arrhythmia during a normal breathing range of 13 breaths per minute). This was completed between the first (guessing) and second (no-guessing) heartbeat tapping conditions. As this tone could be treated as a maximally precise signal, any variations in precision estimates under the model are better explained as due to motor response noise. These “tone precision” estimates could therefore be used as a covariate in analyses to better isolate individual differences not attributable to motor stochasticity in the timing of key presses.

Directly after completing each task condition, individuals were asked the following using a visual analogue scale:

“How intensely did you feel your heartbeat?” or “How intensely did you hear the tone?”

“How accurate was your performance?”

“How difficult was the previous task?”

Each scale had anchors of “not at all” and “extremely” on the two ends. Numerical scores could range from 0 to 100.

### Physiological measurements

The objective timing of participants’ heartbeats was verified throughout the task using cardiac signals from a lead II EKG and pulse waveform signals from a photoplethysmography (PPG) device attached to the ear lobe (via a BIOPAC MP150 acquisition unit). Note, however, that PPG was not attached during the heartbeat tapping task as people can sometimes cutaneously feel their heartbeat through the compression of a PPG clip (28). Response times were collected using a task implemented in PsychoPy, with data collection synchronized via a parallel port interface.

EKG and response data were scored using MATLAB code developed in-house. Each participant’s pulse transit time (PTT) was estimated as the median delay between R wave and the corresponding inflection in the PPG signal.

### Computational model

A detailed description of the computational modeling approach can be found in our prior report (15). All steps described there were carried out identically in the current study. Briefly, each task time series was divided into intervals corresponding to the periods of time directly before and after each heartbeat. Potentially perceivable heartbeats were based on the timing of the peak of the EKG R-wave + 200 milliseconds (ms), based on previous estimates for the PTT (29). The length of each interval in the timeseries (i.e., before vs. after a heartbeat) was based on dividing the time period between every two successive heartbeats (+ 200 ms) in half and then treating the after-beat intervals as the time periods in which the systole (heart muscle contraction) signal was present and in which a key press should be chosen if it was felt. Before-beat intervals were treated as the diastole (heart muscle relaxation) period, in which tapping should not occur. Each interval was treated as a “trial” in which either a tap or no tap could be chosen and in which either a systole or diastole signal was present.

To model perception, we used a hidden Markov model with two time points per “trial”, corresponding, respectively, to a formal trial start state and the presentation of a systole or diastole signal. **Table 2** describes each element of the model in detail (see our prior report for additional motivations and justification of model choices (15)). Heartbeat perception (state inference) depended on a bias in prior beliefs that a heartbeat would be felt (*pHB*) and the precision of the interoceptive sensory signal (*IP*).

**Table 2.**
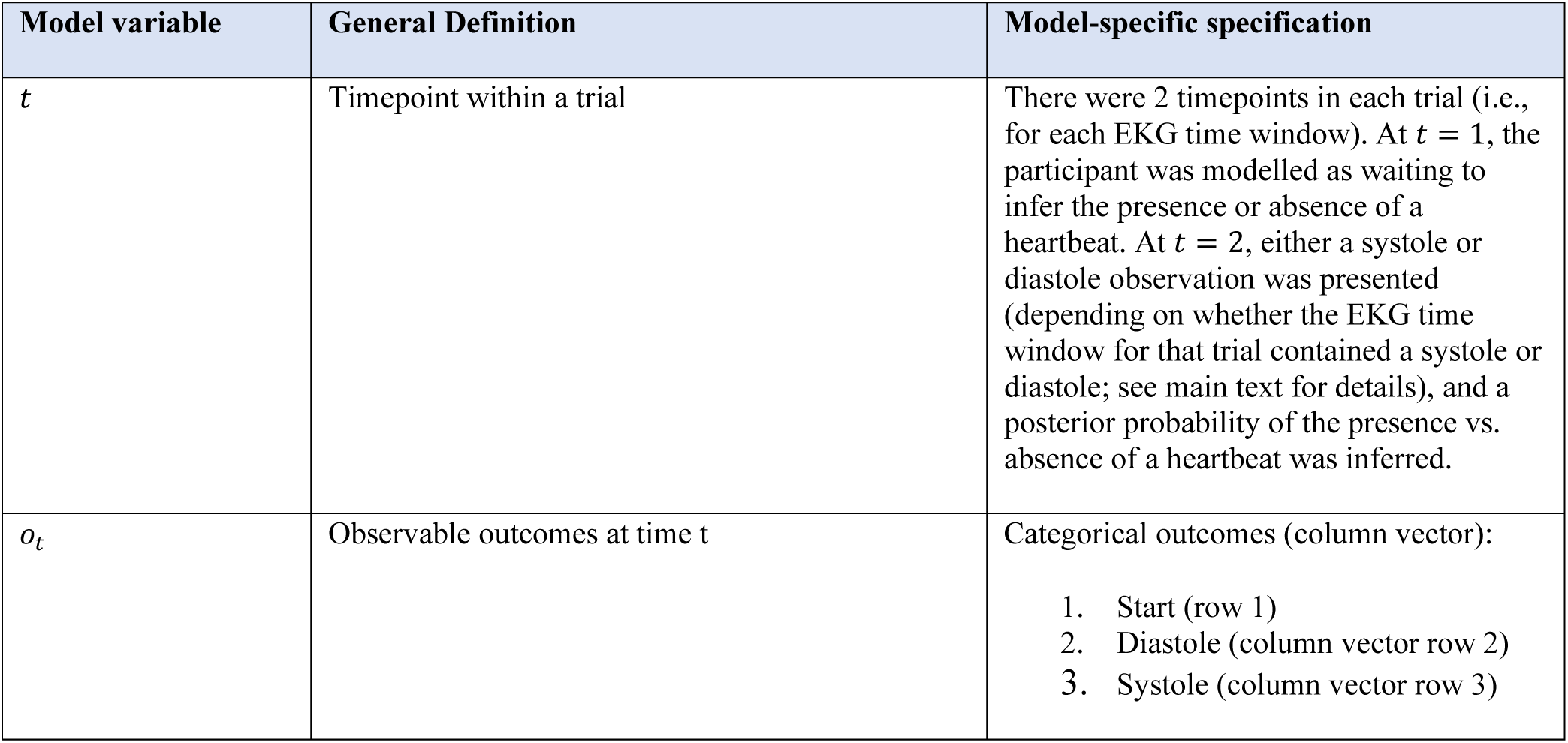
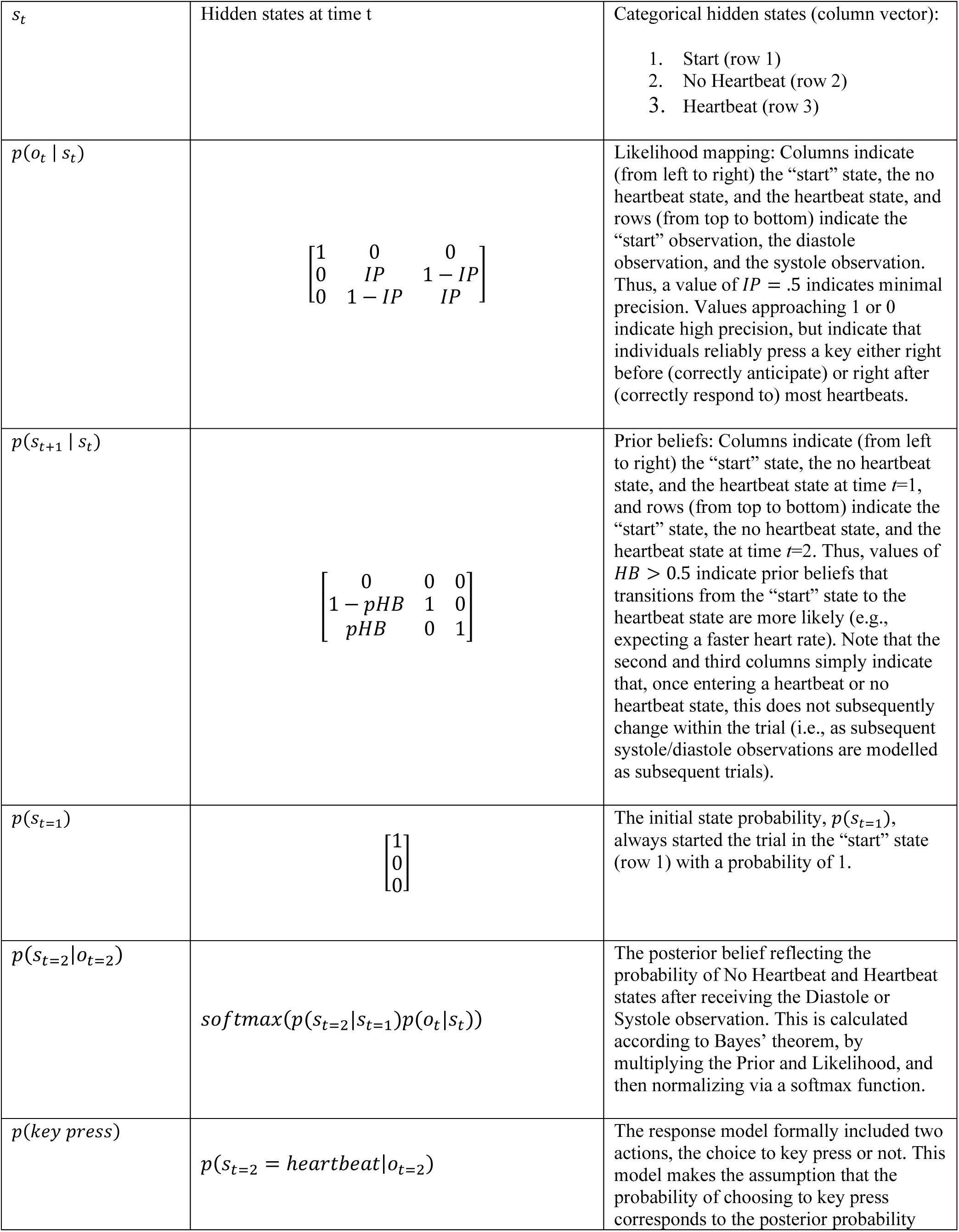
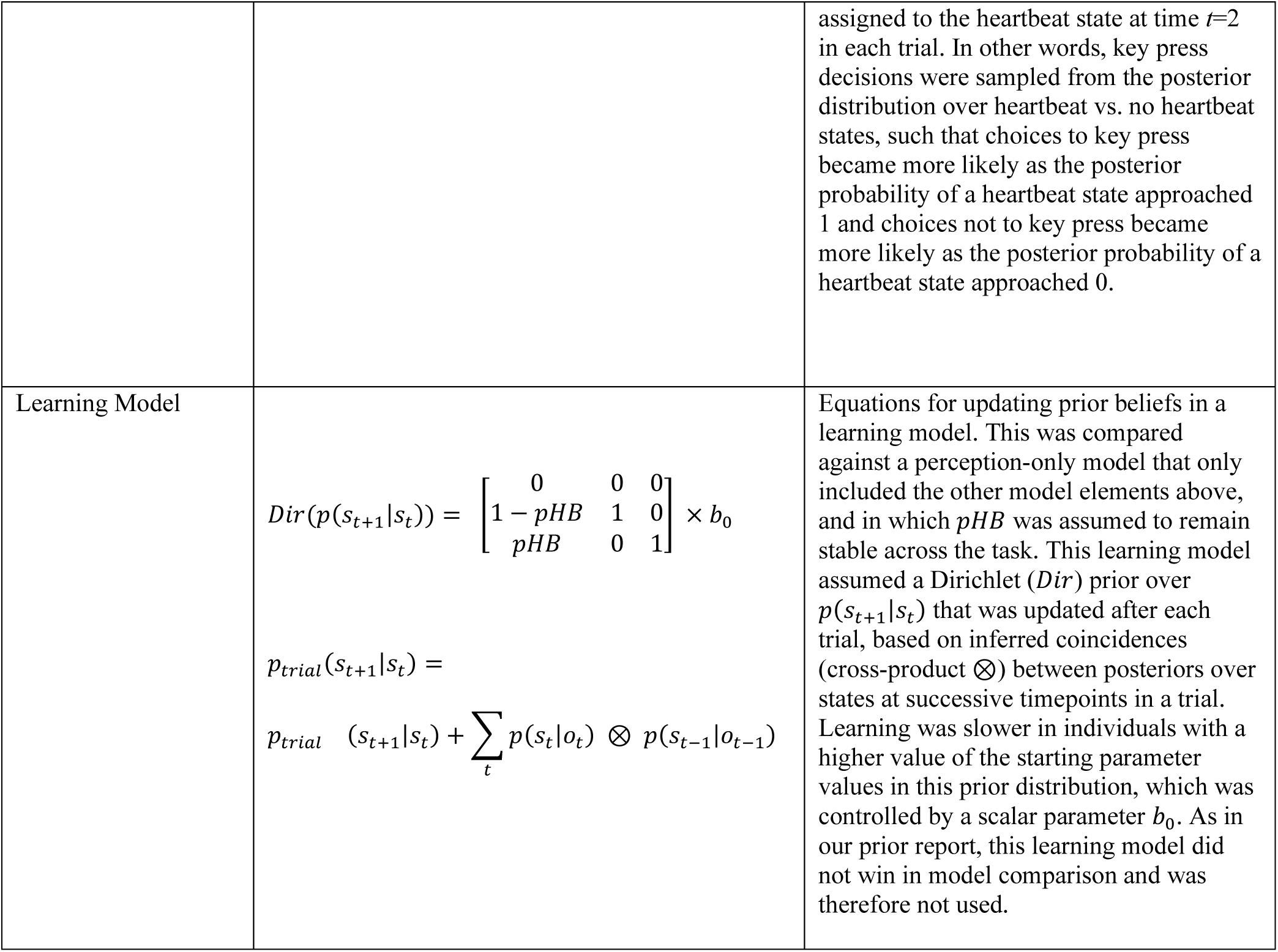
Hidden Markov model of heartbeat tapping task.

| Model variable | General Definition | Model-specific specification |
| --- | --- | --- |
| $t$ | Timepoint within a trial | There were 2 timepoints in each trial (i.e., for each EKG time window). At $t = 1$, the participant was modelled as waiting to infer the presence or absence of a heartbeat. At $t = 2$, either a systole or diastole observation was presented (depending on whether the EKG time window for that trial contained a systole or diastole; see main text for details), and a posterior probability of the presence vs. absence of a heartbeat was inferred. |
| $o_t$ | Observable outcomes at time $t$ | Categorical outcomes (column vector):<br>1. Start (row 1)<br>2. Diastole (column vector row 2)<br>3. Systole (column vector row 3) |
| $s_t$ | Hidden states at time t | Categorical hidden states (column vector):<br>1. Start (row 1)<br>2. No Heartbeat (row 2)<br>3. Heartbeat (row 3) |
| $p(o_t \vert s_t)$ | $\begin{bmatrix} 1 & 0 & 0 \\ 0 & IP & 1 - IP \\ 0 & 1 - IP & IP \end{bmatrix}$ | Likelihood mapping: Columns indicate (from left to right) the “start” state, the no heartbeat state, and the heartbeat state, and rows (from top to bottom) indicate the “start” observation, the diastole observation, and the systole observation. Thus, a value of $IP = .5$ indicates minimal precision. Values approaching 1 or 0 indicate high precision, but indicate that individuals reliably press a key either right before (correctly anticipate) or right after (correctly respond to) most heartbeats. |
| $p(s_{t+1} \vert s_t)$ | $\begin{bmatrix} 0 & 0 & 0 \\ 1 - pHB & 1 & 0 \\ pHB & 0 & 1 \end{bmatrix}$ | Prior beliefs: Columns indicate (from left to right) the “start” state, the no heartbeat state, and the heartbeat state at time $t=1$, and rows (from top to bottom) indicate the “start” state, the no heartbeat state, and the heartbeat state at time $t=2$. Thus, values of $HB > 0.5$ indicate prior beliefs that transitions from the “start” state to the heartbeat state are more likely (e.g., expecting a faster heart rate). Note that the second and third columns simply indicate that, once entering a heartbeat or no heartbeat state, this does not subsequently change within the trial (i.e., as subsequent systole/diastole observations are modelled as subsequent trials). |
| $p(s_{t=1})$ | $\begin{bmatrix} 1 \\ 0 \\ 0 \end{bmatrix}$ | The initial state probability, $p(s_{t=1})$, always started the trial in the “start” state (row 1) with a probability of 1. |
| $p(s_{t=2}\vert o_{t=2})$ | $softmax(p(s_{t=2}\vert s_{t=1})p(o_t\vert s_t))$ | The posterior belief reflecting the probability of No Heartbeat and Heartbeat states after receiving the Diastole or Systole observation. This is calculated according to Bayes’ theorem, by multiplying the Prior and Likelihood, and then normalizing via a softmax function. |
| $p(key\ press)$ | $p(s_{t=2} = heartbeat\vert o_{t=2})$ | The response model formally included two actions, the choice to key press or not. This model makes the assumption that the probability of choosing to key press corresponds to the posterior probability |
| | | assigned to the heartbeat state at time $t=2$ in each trial. In other words, key press decisions were sampled from the posterior distribution over heartbeat vs. no heartbeat states, such that choices to key press became more likely as the posterior probability of a heartbeat state approached 1 and choices not to key press became more likely as the posterior probability of a heartbeat state approached 0. |
| Learning Model | $Dir(p(s_{t+1}\vert s_t)) = \begin{bmatrix} 0 & 0 & 0 \\ 1 - pHB & 1 & 0 \\ pHB & 0 & 1 \end{bmatrix} \times b_0$ $p_{trial}(s_{t+1}\vert s_t) =$ $p_{trial}(s_{t+1}\vert s_t) + \sum_t p(s_t\vert o_t) \otimes p(s_{t-1}\vert o_{t-1})$ | Equations for updating prior beliefs in a learning model. This was compared against a perception-only model that only included the other model elements above, and in which $pHB$ was assumed to remain stable across the task. This learning model assumed a Dirichlet ($Dir$) prior over $p(s_{t+1}\vert s_t)$ that was updated after each trial, based on inferred coincidences (cross-product $\otimes$) between posteriors over states at successive timepoints in a trial. Learning was slower in individuals with a higher value of the starting parameter values in this prior distribution, which was controlled by a scalar parameter $b_0$. As in our prior report, this learning model did not win in model comparison and was therefore not used. |

As in our original study, we also assessed evidence for a learning model (see **Table 2**) through Bayesian model comparison with a perception-only model in which prior beliefs did not change over time. The learning model assumed prior beliefs, *p*(*s*_t+1_|*s*_t_), could update after each new observation, and that learning could happen at different rates for each individual based on an inverse scalar parameter *b*_O_ (i.e., higher values indicate slower learning).

Thus, the final parameters estimated for each participant included the *IP*, *pHB*, and *b*_O_. The parameter values that maximized the likelihood of each participant’s responses were identified using variational Laplace (30), implemented within the *spm_nlsi_Newton.m* parameter estimation routine available within the freely available SPM12 software package (Wellcome Trust Centre for Neuroimaging, London, UK, http://www.fil.ion.ucl.ac.uk/spm). The same prior means (*IP* = .5, *pHB* = .5, and *b*_O_ = 1) and variances (.5) for estimation were set as in our initial study.

After estimation, the “raw” *IP* parameter values (*IP*_raw_) were transformed to capture two distinct constructs of interest. As described in **Table 2**, because *IP*_raw_ values both above and below .5 indicate higher precision (i.e., values below approaching 0 indicate reliable anticipatory key presses, whereas values approaching 1 indicate reliable key presses after each systole), our ultimate measure of precision was recalculated by centering *IP*_raw_ on 0 and taking its absolute value as follows:

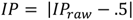

This means that *IP* has a minimum value of 0 and a maximum value of 0.5. The *IP*_raw_ values were then instead used to assess individual differences in the tendency to key press in an anticipatory or reactive (*AvR*) fashion:

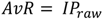

Higher *AvR* values (> 0.5) thus indicated a stronger tendency to reactively key press in response to a systole as opposed to key pressing in an anticipatory fashion (< 0.5).

### Quality Control and Final Sample Sizes

Prior to performing our analyses, several participants were removed due to quality control checks. Specifically, 172 individuals were removed due to “cheating” (i.e., review of their video recording revealed that they took their pulse while performing the task), and 22 individuals were removed due to being outliers when performing the tone task (using Iterative Grubb’s with *p* < 0.01) – assumed to reflect inappropriate engagement during the task (e.g., being inattentive, tapping rapidly without listening to the tones, etc.). This resulted in 285 participants, including 59 HCs, 4 iANX, 26 iDEP, 88 iDEP+ANX, 84 iSUDs, and 24 iEDs. Due to the small sample size in the anxiety group, these 4 participants were excluded from disorder-specific analyses.

### Statistical analysis

#### Primary replication analyses

Analyses replicated those carried out in our prior study. We first used JZS Bayes factor (BF) analyses (repeated-measures ANOVAs) with default prior scales in R (31) to compare evidence for models with transdiagnostic vs. diagnosis-specific effects. These models included group, condition, and their interaction, and either coded group as two levels (HCs vs. all patients) or 5 levels (HCs, iDEP, iDEP+ANX, iEDs, and iSUDs). This model comparison was done to assess whether the results were better interpreted as transdiagnostic or diagnosis-specific. Based on our previous study, we expected that BF analyses would support a model assuming a single transdiagnostic group. Linear mixed effects models (LMEs) were then conducted (using the lme4 package in R (32)) to confirm significant group differences (i.e., between HCs and either the transdiagnostic group or the four patient groups) for each parameter, and if they differ between conditions (i.e., guessing, no-guessing, breath-hold). To confirm the specificity of significant effects, we also ran models that accounted for individual differences in age, sex, body mass index (BMI), median pulse transit time (PTT), number of heartbeats (and its interaction with group and condition), and medication status (i.e., one analysis per model parameter). Due to missing values in PTT (*n* = 89) and BMI (*n* = 1), the group-level averages were imputed to maximize sample size while not influencing the effect of either predictor in the models. One additional participant was missing BMI data from the time of task completion but had data in the months following. A regression line was fit to the data points available and the best fit value at the time of task completion was imputed for this individual. As in the original study, we also included *IP* estimates for the auditory control condition as a covariate to account for variability in behavior attributable to motor stochasticity (e.g., differences in reaction times). Categorical variables (i.e., group, condition, sex, and medication status) were sum-coded in all models.

#### Secondary analyses

As in our prior study, for comparison to traditional interoception measures we performed identical analyses predicting a commonly-used counting accuracy measure of performance (33). These analyses were also performed for several self-report and physiological variables in each condition, including heart rate and self-reported confidence in performance, task difficulty, and perceived heartbeat intensity. We further tested for relationships between model parameters and these measures, as well as with age, sex, BMI, and PTT. Based on our prior results, we expected positive relationships in the no-guessing and breath-hold conditions between both model parameters and self-reported heartbeat intensity, and a positive relationship between *pHB* and self-reported confidence. In the no-guessing condition, we expected a negative correlation between *pHB* and self-reported difficulty. We also expected weak and strong positive relationships between counting accuracy and *IP* and *pHB*, respectively. Separate ANOVAs and chi-squared analyses also tested for significant differences between groups in other individual difference variables, including: age, sex, PHQ, OASIS, DAST, BMI, PTT, heartrate, number of key presses, and self-reported beliefs about felt heartbeat intensity, task difficulty, and confidence in task performance.

Correlations between model parameters were examined both within and between conditions. Based on our prior report, we expected to see weak positive correlations between both *IP* in the no-guessing and breath-hold conditions, weak positive correlations between *pHB* in the guessing and no-guessing conditions, and moderate positive correlations between *pHB* in the no-guessing and breath-hold conditions. We also expected to find weak positive correlations between *IP* and *pHB* in the no-guessing and breath-hold conditions, and weak negative relationships between *AvR* and both *IP* and *pHB* across all conditions.

We also ran correlational analyses with continuous scores on some of the clinical measures (PHQ, OASIS, DAST) gathered, excluding HCs, to assess whether model parameters provided additional information about symptom severity. Based on our prior exploratory results, we predicted the following relationships in the current dataset: In the no-guessing condition, *IP* would be negatively associated with both depression (PHQ) and anxiety (OASIS) severity, and positively associated with substance use severity (DAST); *pHB* would also be positively associated with substance use severity (DAST). In the breath-hold condition, *pHB* would be positively associated with substance use severity (DAST), and higher anxiety (OASIS) would be associated with a more anticipatory response pattern (*AvR*).

The previous dataset included common self-report measures of interoception, including the Multidimensional Assessment of Interoceptive Awareness (MAIA (34)) the Toronto Alexithymia Scale (TAS-20 (35)), and the Anxiety Sensitivity Index (ASI (36)), which were correlated with model parameters for some subscales in exploratory analyses (see **Supplementary Figure S5** in our prior study). We also attempted to replicate these exploratory results.

#### Sample comparisons and combined sample analyses

We compared the initial exploratory sample with this confirmatory sample on demographic and clinical characteristics as well as model parameter values. In addition to the preregistered replication analyses above, we then combined these two samples (N = 719) to maximize power for examining potential differences between individuals within the smaller diagnostic categories. First, we tested whether model parameters could predict the presence of specific substance use or affective disorders relative to healthy comparisons using separate logistic regressions. Each regression included both parameters (separated by condition) as predictors of diagnostic status (coding 0 = healthy comparison and 1 = those with the specific disorder, excluding all other participants). We also performed these tests predicting disorders without comorbidities when the sample size allowed it. Following this, we performed analogous regressions within groups comparing those with a specific disorder (coded as 1) to those without it (coded as 0) excluding HCs. This allowed us to determine if the model parameters could differentiate disorders from one another. All logistic regressions for a given disorder were performed separately in those with vs. without substance use disorders (SUDs).

Finally, we sought to evaluate if a model including interoceptive precision parameters that was trained on the exploratory sample could successfully classify participants in the confirmatory sample by diagnostic status. To this end, we performed prediction classification analyses (logistic regressions) including the *IP* parameters in each of the three HB tapping conditions. We first tested whether HCs could be separated from all patients, irrespective of diagnosis. We then removed HCs and tested whether we could correctly classify specific diagnoses. Training and class prediction within logistic regressions was carried out using the *statsmodels* package in Python. Frequency balancing of the training dataset was performed using the *skylearn* package. These analyses allowed us to evaluate predictive accuracy based on area-under-the-curve (AUC) scores and confusion matrices in the test dataset.

## Results

### Participant characteristics

Complete information on sample size, demographics, and symptom screening measures is provided in **Table 3**. Initial t-tests indicated that the iED were younger than all other groups (*p* ≤ .05) and that iDEP+ANX were younger than iDEP and iSUD (*p* ≤ .042). Additionally, the iED had significantly lower BMI than all other groups (*p* ≤ .032). No other groups differed significantly. A chi-squared analysis also showed significant differences in the proportion of males to females between groups (chi-squared = 18.00, df = 5, *p* = .003) specifically between HCs and iED (*p* = .006), HCs and iDEP+ANX (*p* = .007), and iDEP and iDEP+ANX (*p* = .046). While not included in the table below, some participants in our clinical groups were taking psychotropic medications at the time of data collection (25% iANX, 42% iDEP, 27% iDEP+ANX, 50% iED, and 49% iSUD). Therefore, in our analyses of model parameters, we also confirm our results after controlling for these other factors.

**Table 3.**
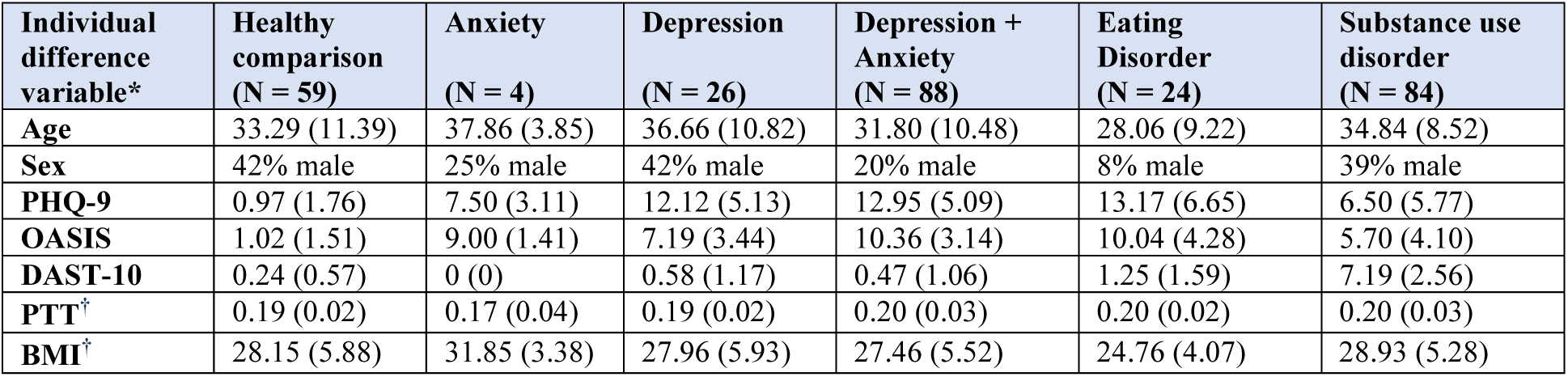
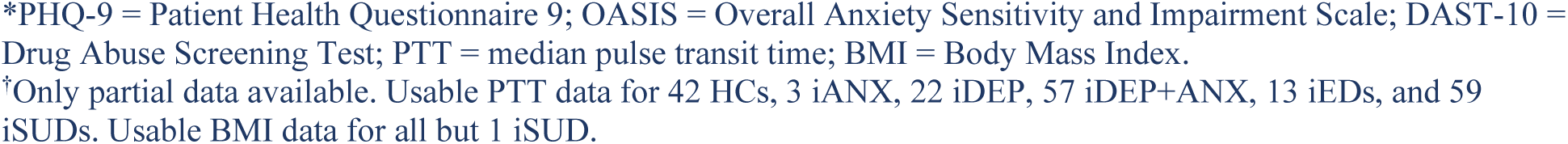
Mean (and standard deviation) for clinical and demographic variables.

| Individual difference variable* | Healthy comparison (N = 59) | Anxiety (N = 4) | Depression (N = 26) | Depression + Anxiety (N = 88) | Eating Disorder (N = 24) | Substance use disorder (N = 84) |
| --- | --- | --- | --- | --- | --- | --- |
| Age | 33.29 (11.39) | 37.86 (3.85) | 36.66 (10.82) | 31.80 (10.48) | 28.06 (9.22) | 34.84 (8.52) |
| Sex | 42% male | 25% male | 42% male | 20% male | 8% male | 39% male |
| PHQ-9 | 0.97 (1.76) | 7.50 (3.11) | 12.12 (5.13) | 12.95 (5.09) | 13.17 (6.65) | 6.50 (5.77) |
| OASIS | 1.02 (1.51) | 9.00 (1.41) | 7.19 (3.44) | 10.36 (3.14) | 10.04 (4.28) | 5.70 (4.10) |
| DAST-10 | 0.24 (0.57) | 0 (0) | 0.58 (1.17) | 0.47 (1.06) | 1.25 (1.59) | 7.19 (2.56) |
| PTT<sup>†</sup> | 0.19 (0.02) | 0.17 (0.04) | 0.19 (0.02) | 0.20 (0.03) | 0.20 (0.02) | 0.20 (0.03) |
| BMI<sup>†</sup> | 28.15 (5.88) | 31.85 (3.38) | 27.96 (5.93) | 27.46 (5.52) | 24.76 (4.07) | 28.93 (5.28) |

\*PHQ-9 = Patient Health Questionnaire 9; OASIS = Overall Anxiety Sensitivity and Impairment Scale; DAST-10 = Drug Abuse Screening Test; PTT = median pulse transit time; BMI = Body Mass Index.
†Only partial data available. Usable PTT data for 42 HCs, 3 iANX, 22 iDEP, 57 iDEP+ANX, 13 iEDs, and 59 iSUDs. Usable BMI data for all but 1 iSUD.

### Sample Comparison

Bayesian t-tests comparing clinical and demographic variables between the previous and current samples (by group) are reported in **Supplemental Table S1.** There were no significant differences (BFs ≤ 2.01) apart from BMI in the iEDs, which was higher in the confirmatory sample than the exploratory sample (BF = 3.87). **Figure 1** below shows the diagnostic breakdowns of the current and previous samples.

**Figure 1.**
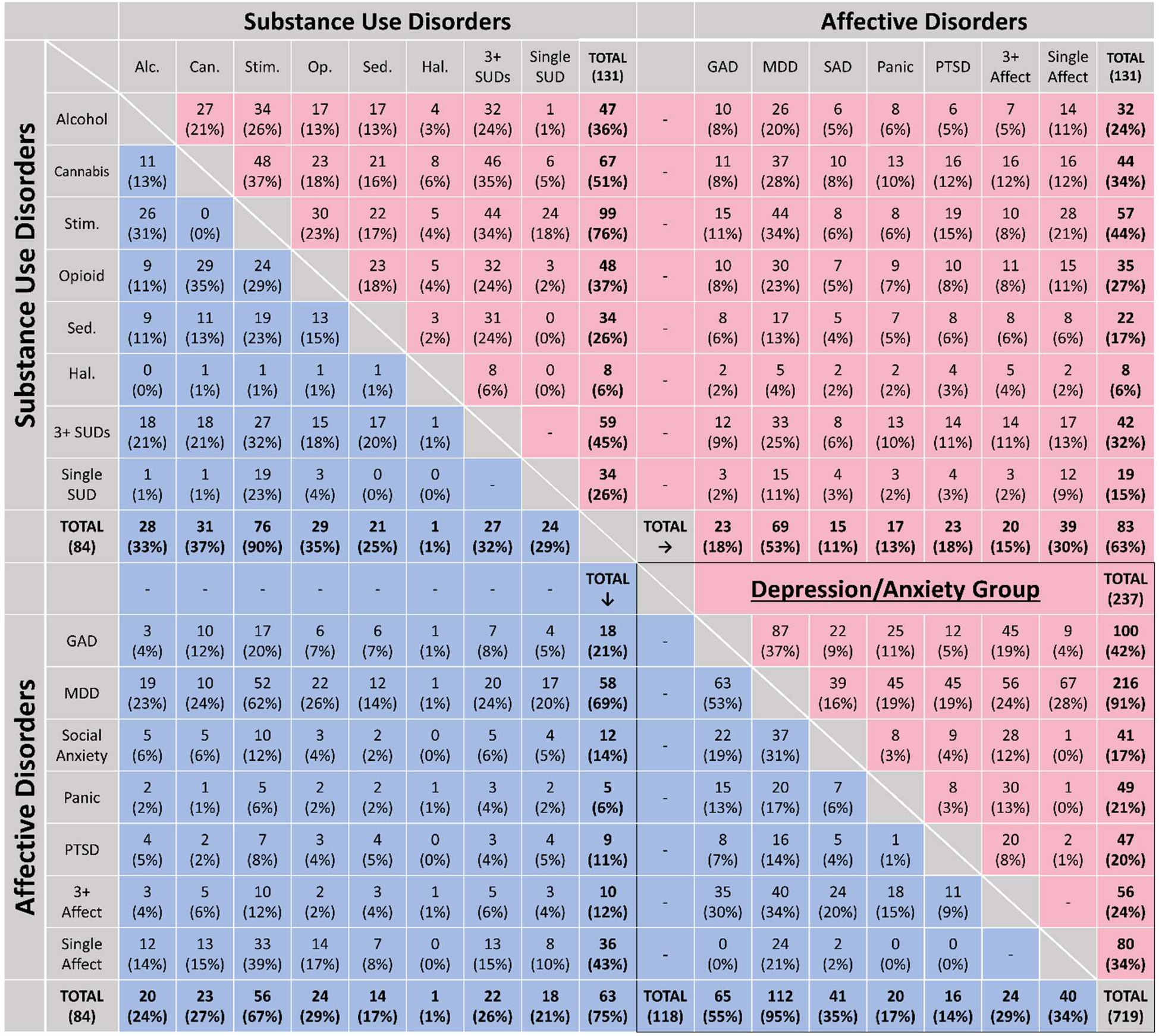
DSM-IV/DSM-5 psychiatric diagnosis composition within exploratory (above diagonal) and confirmatory (below diagonal) samples. Two individuals (both in the exploratory sample) were given a diagnosis of polysubstance use disorder without further specification and were included in the category of those with 3+ SUDs. Alc. = alcohol use disorder; Can. = cannabis use disorder; GAD = generalized anxiety disorder; Hal. = hallucinogen use disorder; MDD = major depressive disorder; Op. = opioid use disorder; PTSD = posttraumatic stress disorder; SAD = social anxiety disorder; Sed. = sedative use disorder; Stim. = stimulant use disorder; SUD = substance use disorder.

### Interoceptive Perturbation Validation

Task-related self-report variables by group are shown in **Table 4** split by condition (Panels A-D). As expected, across all participants self-reported heartbeat intensity, confidence in task performance, and task difficulty differed significantly between the three heartbeat tapping conditions in separate LMEs (intensity: *F*(2,567) = 58.99, *p* < .001; confidence: *F*(2,568) = 10.40, *p* < .001; difficulty: *F*(2,568) = 17.49, *p* < .001), reflecting a greater perceived intensity of heartbeat sensations and greater confidence during the breath-hold perturbation than in the other 2 conditions, as well as lower difficulty in the breath-hold condition than in the no-guessing condition (and greater difficulty in the no-guessing than guessing condition; *p* < .001 for all post-hoc comparisons). An LME analysis of heart rate revealed a significant difference in the number of heartbeats between conditions (*F*(2,568) = 4.11, *p* = .017), reflecting a faster heart rate in the breath-hold condition than in the no-guessing condition (*p* = .005). This effect did not differ by group.

**Table 4.** Summary statistics for all task-related variables.

| **<u>Guessing Condition</u>** | **Healthy Comparisons (N = 59)** | **Anxiety (N = 4)** | **Depression (N = 26)** | **Depression + Anxiety (N = 88)** | **Eating Disorder (N = 22)** | **Substance Use Disorder (N = 84)** | **p-value\*** |
| --- | --- | --- | --- | --- | --- | --- | --- |
| **Number of taps** | 60.41<br>(24.64) | 62.5<br>(49.62) | 54.69<br>(34.79) | 64.63<br>(30.49) | 59.04<br>(25.32) | 74.95<br>(51.32) | .095 |
| **Number of heartbeats** | 66.81<br>(11.60) | 69.75<br>(2.5) | 55.31<br>(11.07) | 59.51<br>(15.13) | 60.88<br>(22.18) | 49.71<br>(15.31) | .345 |
| **Self-reported difficulty** | 58.86<br>(25.42) | 77.50<br>(1.73) | 55.31<br>(29.42) | 59.51<br>(25.07) | 60.88<br>(27.12) | 49.71<br>(28.45) | .072 |
| **Self-reported confidence** | 38.76<br>(23.87) | 28.00<br>(6.83) | 37.77<br>(18.63) | 33.83<br>(28.89) | 26.13<br>(18.35) | 38.75<br>(22.74) | .116 |
| **Self-reported intensity** | 29.46<br>(22.20) | 16.00<br>(23.04) | 25.81<br>(22.25) | 24.59<br>(20.99) | 17.54<br>(17.08) | 28.46<br>(23.56) | .202 |
| **Counting accuracy** | 0.74<br>(0.24) | 0.51<br>(0.39) | 0.59<br>(0.29) | 0.62<br>(0.61) | 0.70<br>(0.25) | 0.45<br>(0.95) | .173 |

\*p-values correspond to the results of ANOVAs comparing the groups (i.e., not including all task conditions within the analysis as reported in the main text).

Panel B.
| **<u>No-Guessing Condition</u>** | **Healthy Comparisons**<br>**(N = 59)** | **Anxiety**<br>**(N = 4)** | **Depression**<br>**(N = 26)** | **Depression + Anxiety**<br>**(N = 88)** | **Eating Disorder**<br>**(N = 22)** | **Substance Use Disorder**<br>**(N = 83)** | **p-value** |
| --- | --- | --- | --- | --- | --- | --- | --- |
| **Number of taps** | 24.34<br>(26.79) | 15.50<br>(14.82) | 21.46<br>(22.17) | 21.15<br>(24.42) | 24.88<br>(21.32) | 24.98<br>(26.97) | .885 |
| **Number of heartbeats** | 65.59<br>(10.86) | 68.50<br>(2.38) | 67.15<br>(14.39) | 69.44<br>(16.32) | 63.46<br>(22.68) | 66.86<br>(17.89) | .610 |
| **Self-reported difficulty** | 65.85<br>(29.42) | 70.75<br>(39.23) | 63.96<br>(34.39) | 68.06<br>(24.00) | 61.21<br>(28.90) | 62.02<br>(29.18) | .757 |
| **Self-reported confidence** | 38.24<br>(28.38) | 27.25<br>(19.26) | 27.85<br>(29.71) | 39.77<br>(28.85) | 29.33<br>(24.54) | 39.77<br>(28.71) | .213 |
| **Self-reported intensity** | 22.80<br>(23.36) | 14.50<br>(23.69) | 23.73<br>(25.04) | 22.68<br>(23.46) | 26.21<br>(23.43) | 24.21<br>(21.73) | .946 |
| **Counting accuracy** | 0.31<br>(0.29) | 0.22<br>(0.20) | 0.31<br>(0.29) | 0.10<br>(1.71) | 0.37<br>(0.33) | 0.32<br>(0.33) | .683 |

Panel C.
| **<u>Breath Hold Condition</u>** | **Healthy Comparisons**<br>**(N = 59)** | **Anxiety**<br>**(N = 4)** | **Depression**<br>**(N = 26)** | **Depression + Anxiety**<br>**(N = 87)** | **Eating Disorder**<br>**(N = 24)** | **Substance Use Disorder**<br>**(N = 82)** | **p-value** |
| --- | --- | --- | --- | --- | --- | --- | --- |
| **Number of taps** | 29.47<br>(26.97) | 17.75<br>(15.44) | 23.81<br>(24.26) | 24.16<br>(22.49) | 23.04<br>(22.13) | 24.39<br>(22.35) | .711 |
| **Number of heartbeats** | 69.86<br>(13.00) | 72.50<br>(3.87) | 71.04<br>(11.32) | 69.35<br>(17.39) | 68.88<br>(11.11) | 68.52<br>(20.51) | .983 |
| **Self-reported difficulty** | 51.14<br>(28.14) | 47.50<br>(35.01) | 51.96<br>(32.50) | 57.84<br>(27.33) | 61.17<br>(24.07) | 54.68<br>(28.01) | .582 |
| **Self-reported confidence** | 48.05<br>(26.97) | 22.25<br>(6.24) | 32.69<br>(27.24) | 43.49<br>(25.00) | 31.29<br>(18.41) | 47.68<br>(26.98) | **.006** |
| **Self-reported intensity** | 42.61<br>(29.81) | 36.50<br>(22.99) | 37.69<br>(30.64) | 38.51<br>(26.06) | 33.87<br>(27.89) | 38.76<br>(28.66) | .869 |
| **Counting accuracy** | 0.37<br>(0.30) | 0.25<br>(0.22) | 0.33<br>(0.33) | 0.27<br>(0.48) | 0.34<br>(0.31) | 0.32<br>(0.31) | .648 |

Panel D.
| <u>Tone Condition</u> | Healthy Comparisons (N = 59) | Anxiety (N = 4) | Depression (N = 26) | Depression + Anxiety (N = 88) | Eating Disorder (N = 24) | Substance Use Disorder (N = 84) | p-value |
| --- | --- | --- | --- | --- | --- | --- | --- |
| Number of taps | 77.73 (0.74) | 78.00 (0.00) | 77.77 (0.65) | 78.10 (0.84) | 78.17 (0.38) | 78.10 (0.87) | .024* |
| Number of heartbeats | 67.75 (11.31) | 71.00 (3.56) | 70.08 (11.19) | 71.35 (15.77) | 64.21 (22.83) | 68.50 (15.74) | .399 |
| Self-reported difficulty | 25.54 (21.77) | 25.25 (18.89) | 22.85 (23.76) | 28.01 (22.98) | 22.29 (16.29) | 28.15 (24.94) | .790 |
| Self-reported confidence | 76.54 (21.13) | 84.00 (10.92) | 76.23 (18.05) | 71.98 (18.38) | 76.75 (14.76) | 70.67 (20.95) | .297 |
| Self-reported intensity | 84.68 (17.41) | 88.00 (7.87) | 82.54 (17.96) | 82.32 (16.60) | 85.79 (13.36) | 78.14 (18.71) | .201 |
| Counting accuracy | 1.00 (0.01) | 1.00 (0) | 1.00 (0.01) | 0.99 (0.01) | 1.00 (0) | 0.99 (0.01) | .310 |

\* Note that, while significant, this reflects differences of less than one tap.

### Bayesian Model Comparison

Similar to our prior report, when comparing models (based on (37, 38)), there was more evidence for the “perception only” model than for the model that included learning prior beliefs for the no-guessing and breath-hold conditions (protected exceedance probability = 1), whereas there was not clear evidence favoring a single model for the guessing condition (protected exceedance probabilities = 0.42 vs. 0.58, slightly favoring the learning model). The learning model was favored in the tone condition (protected exceedance probability = 1). No group differences were observed when comparing model fits between groups.

For consistency/comparability, we use the “perception only” model parameters to compare conditions in our analyses below (as in our previous study), as this model best explained heartbeat tapping behavior overall. The accuracy of this model – defined as the percentage of choices to key press that matched the highest probability action in the model (e.g., a key press occurring when the highest probability percept in the model was a heartbeat) – was 77% across all conditions; by condition, model accuracy was: guessing condition = 67% (SD = 12%); tone condition = 68% (SD = 12%); no-guessing condition = 87% (SD = 13%); breath-hold condition = 85% (SD = 12%).

### Relationship between parameters

Parameter values for each group and condition are listed in **Table 5** (for associated plots, see **Supplementary Figure S1**). Across conditions, all parameters were normally distributed (skew < 2). **Figure 2** shows inter-correlations for each parameter across conditions. As can be seen there, no significant correlations between *IP* across task conditions were found. Correlations between *pHB* estimates across conditions were moderate to strong, most notably between the no-guessing and breath-hold conditions (which also includes the no-guessing instruction). The tendency to tap in an anticipatory vs. reactionary manner (*AvR*) showed a positive relationship between the guessing and breath-hold conditions.

**Figure 2.**
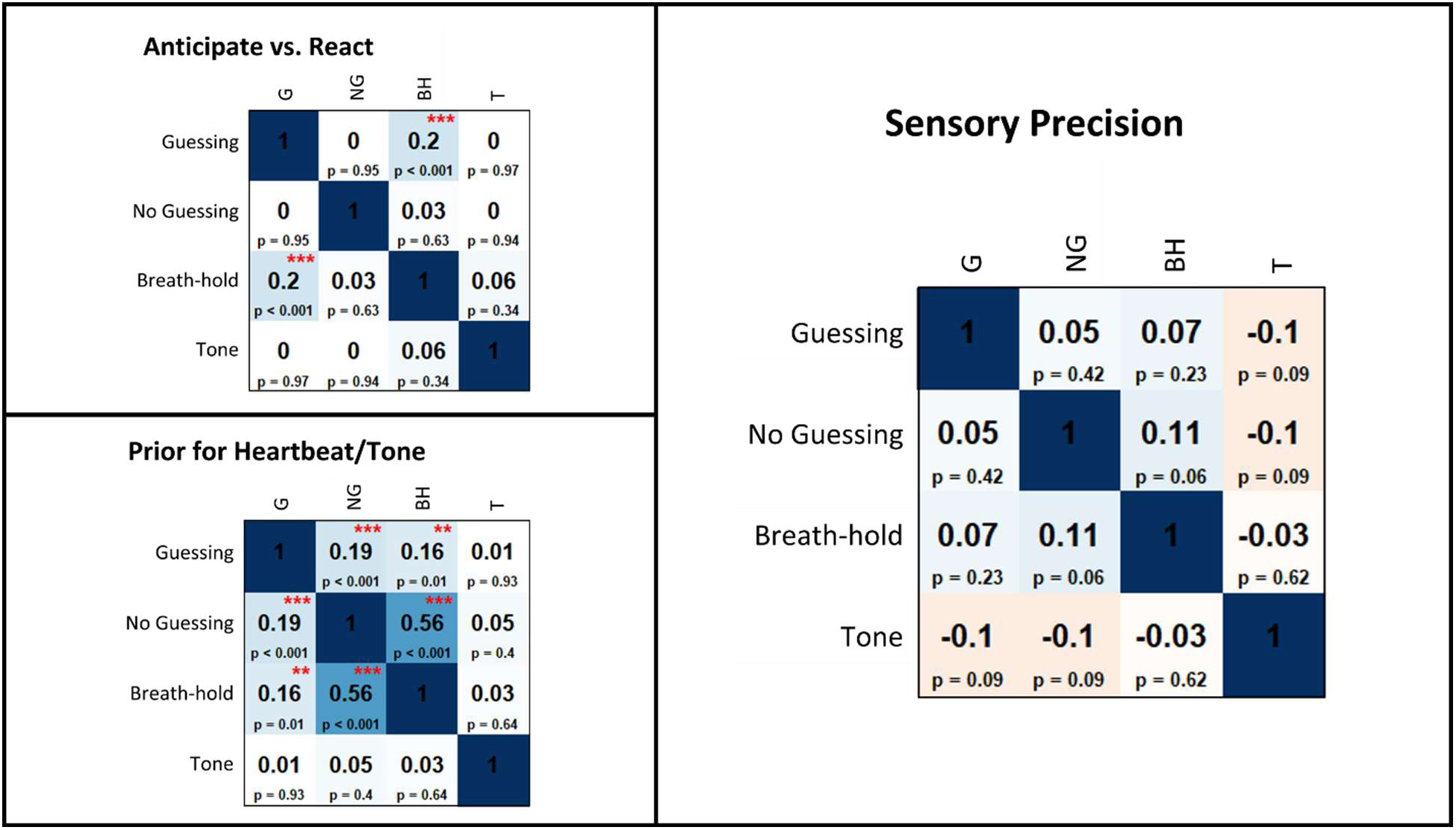
Pearson correlations between model parameters across task conditions across all participants. For reference, correlations at p ≤ .01 or .001 (uncorrected) are marked with two or three red asterisks, respectively.

**Table 5.** Mean (and standard deviation) for model parameters by group and condition.

| Individual difference variable | Healthy Comparisons | Anxiety | Depression | Depression + Anxiety | Eating Disorder | Substance Use Disorder |
| --- | --- | --- | --- | --- | --- | --- |
| **Sensory precision** |  |  |  |  |  |  |
| Guessing | 0.04 (0.04) | 0.05 (0.04) | 0.03 (0.03) | 0.04 (0.05) | 0.05 (0.06) | 0.04 (0.04) |
| No-Guessing | 0.04 (0.05) | 0.03 (0.02) | 0.03 (0.04) | 0.04 (0.04) | 0.04 (0.07) | 0.04 (0.05) |
| Breath-hold | 0.06 (0.07) | 0.04 (0.03) | 0.04 (0.07) | 0.04 (0.04) | 0.04 (0.05) | 0.04 (0.04) |
| Tone | 0.18 (0.14) | 0.23 (0.11) | 0.12 (0.10) | 0.15 (0.12) | 0.23 (0.09) | 0.17 (0.11) |
| **Prior beliefs** |  |  |  |  |  |  |
| Guessing | 0.39 (0.13) | 0.37 (0.24) | 0.35 (0.18) | 0.39 (0.16) | 0.39 (0.16) | 0.43 (0.18) |
| No-Guessing | 0.18 (0.15) | 0.12 (0.08) | 0.16 (0.12) | 0.16 (0.14) | 0.18 (0.13) | 0.18 (0.14) |
| Breath-hold | 0.19 (0.14) | 0.13 (0.08) | 0.16 (0.13) | 0.17 (0.12) | 0.17 (0.12) | 0.17 (0.12) |
| Tone | 0.50 (0.01) | 0.50 (0.01) | 0.50 (0.00) | 0.50 (0.01) | 0.50 (0.01) | 0.50 (0.01) |
| **Anticipate vs. React** |  |  |  |  |  |  |
| Guessing | 0.50 (0.06) | 0.47 (0.06) | 0.50 (0.04) | 0.50 (0.06) | 0.50 (0.08) | 0.49 (0.05) |
| No-Guessing | 0.50 (0.06) | 0.47 (0.02) | 0.50 (0.05) | 0.50 (0.06) | 0.50 (0.08) | 0.51 (0.06) |
| Breath-hold | 0.51 (0.09) | 0.54 (0.04) | 0.48 (0.08) | 0.51 (0.06) | 0.50 (0.07) | 0.50 (0.06) |
| Tone | 0.55 (0.22) | 0.49 (0.29) | 0.54 (0.15) | 0.46 (0.19) | 0.36 (0.21) | 0.44 (0.19) |

**Supplementary Figure S2** shows correlations between parameters across conditions. Most notably, significant correlations were present between *IP* in the no-guessing condition and both *pHB* in the no-guessing (*r* = .25, *p* < .001) and breath-hold conditions (*r* = .26, *p* < .001).

### Parameter face validity

Figure 3 shows the correlations, including some significant relationships (*p* ≤ .05, uncorrected), across all participants between model parameters in each condition and several task-relevant variables. *IP* showed positive relationships with self-reported heartbeat intensity ratings in the no-guessing and breath hold conditions. Additionally, *pHB* was lower in those self-reporting greater difficulty in the no-guessing condition, and higher in those self-reporting higher confidence and higher heartbeat intensity in the breath-hold and guessing conditions. For *IP* in all heartbeat conditions, and for *pHB* in the breath-hold condition, parameters also related to the traditional counting accuracy measure as expected (33). Unlike in the original sample, these results showed that *pHB* was (weakly) negatively correlated with counting accuracy in the guessing condition. The sex difference in *IP* in the breath hold condition reported in our previous report did not replicate in this sample (*t*(280) = -1.09, *p* = .276).

**Figure 3.**
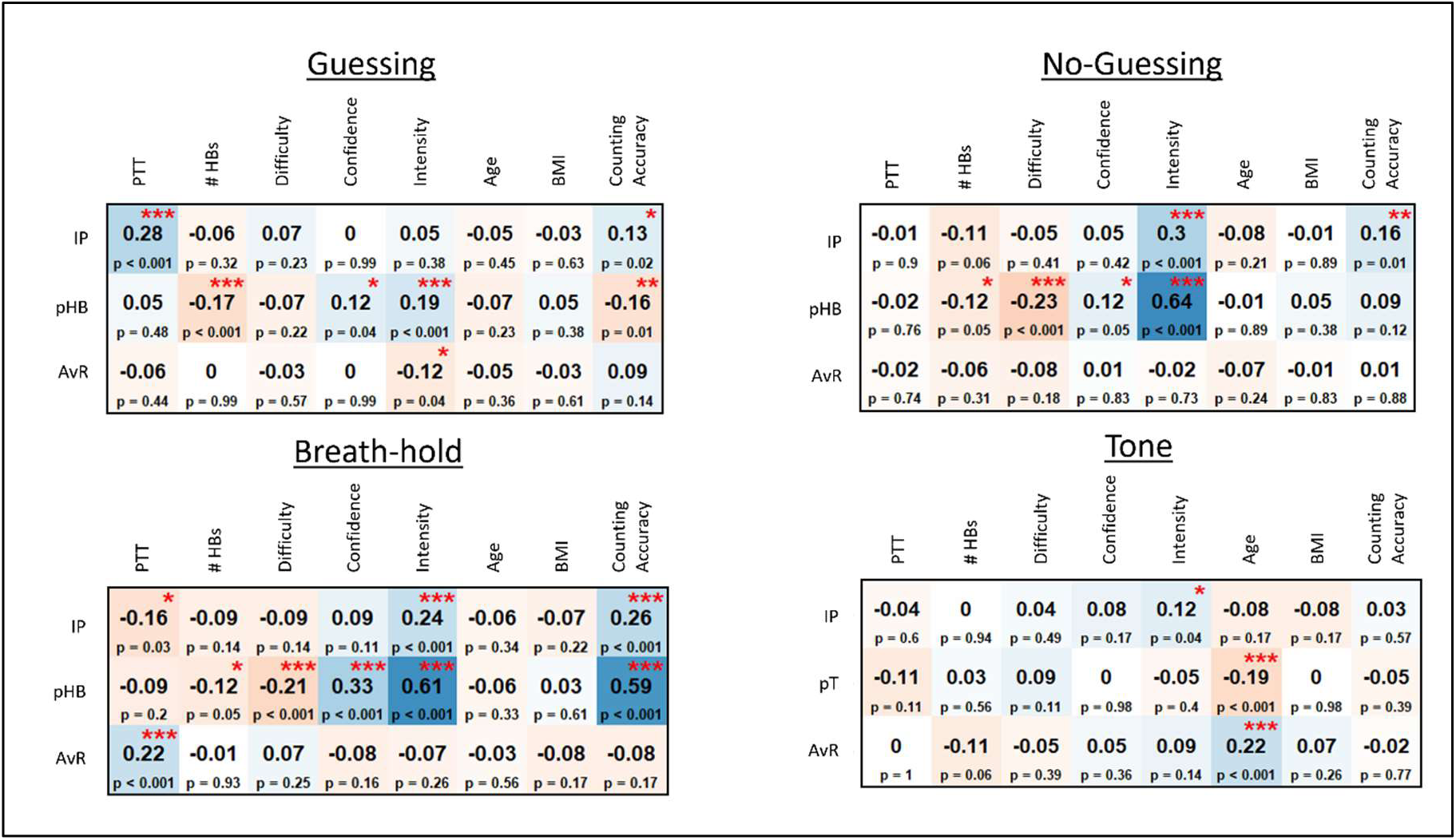
Exploratory Pearson correlations between model parameters and self-report and other task-relevant variables for each task condition across all participants. IP = interoceptive precision parameter, pHB = prior belief for heartbeat parameter, pT = prior belief for tone parameter, AvR = anticipate vs. react strategy parameter, PTT = median pulse transit time, #HBs = number of heartbeats during the task condition, BMI = body mass index. For reference, significant correlations are marked with red asterisks: *p < .05, **p < .01, ***p < .001.

### Group differences

#### Comparison to previous sample

Figure 4 compares the values for *IP* and *pHB* in each condition within the previous and current datasets, separately for HCs and the combined patient group. For an analogous plot of *AvR*, see **Supplemental Figure S3**. All parameters in the tone condition are shown in **Supplemental Figure S4**. As can be seen there, values for each parameter were comparable. Before performing pre-registered analyses below, we tested for differences between our previous and current samples. Bayes factor analyses provided evidence against the presence of differences between samples in all cases (*IP*: BFs between .09 and .33; *pHB*: BFs between .10 and .69). Separate LMs for each condition that included transdiagnostic group, sample, and a group by sample interaction also showed a strong group effect only in the breath-hold condition for *IP* (*F*(1,705) = 26.94, *p* < .001) and no other significant effects (*F* < 2.56, *p* > .110 in all cases).

**Figure 4.**
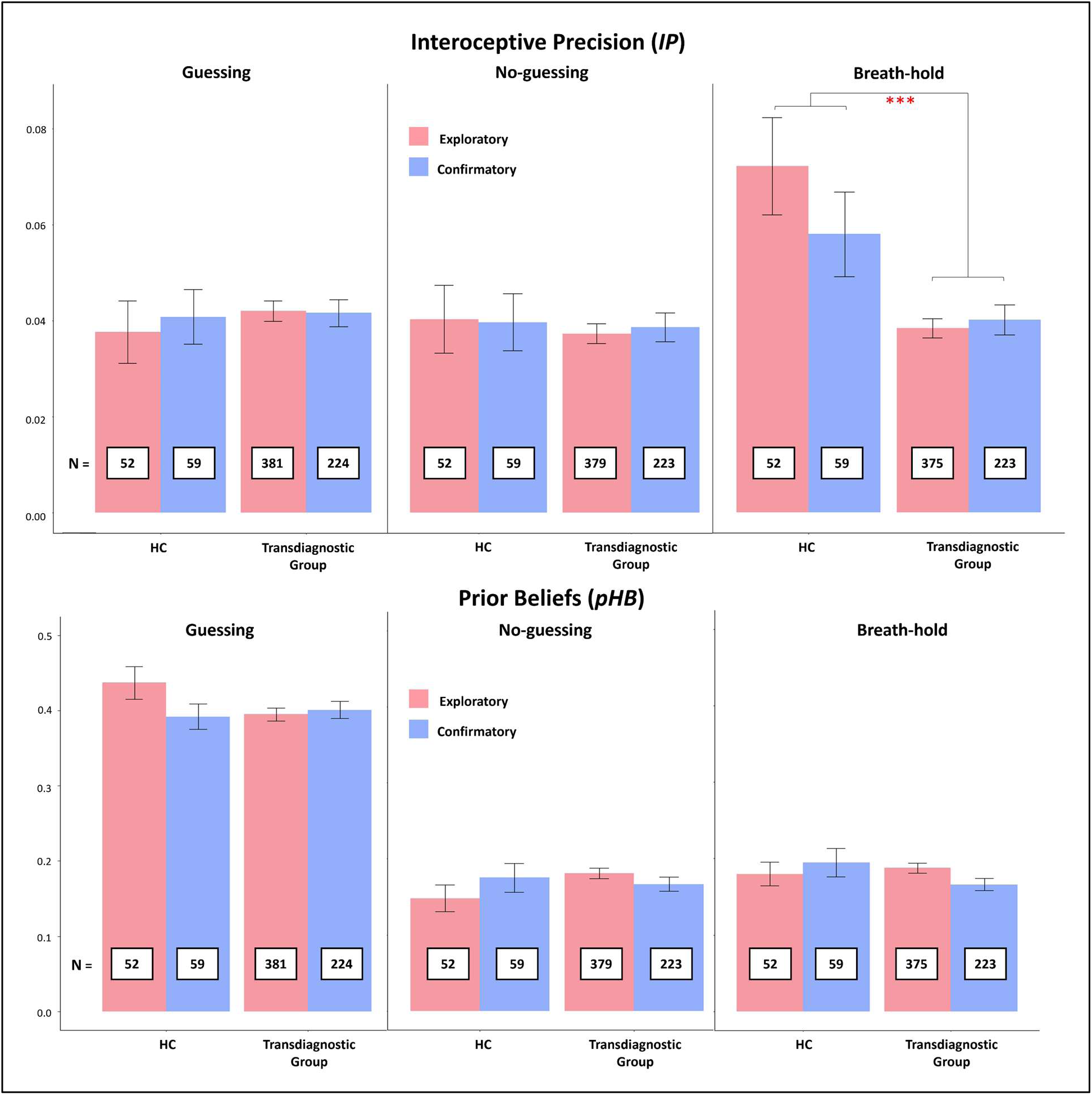
**Top**: Interoceptive precision (*IP*) estimate comparison between the previous (i.e., exploratory) and current (i.e., confirmatory) samples when grouping participants transdiagnostically. Red asterisks indicate a significant difference (\*\*\**p* < .001) in *IP* estimates within an LME between groups in the breath-hold condition (but no difference between samples in either group). Bayes factor analyses also provided moderate to strong evidence supporting an absence of differences between samples in all cases: BFs between .09 (Guessing Condition, Patients) and .33 (Breath-hold Condition, HCs). **Bottom**: Prior expectation (*pHB*) estimate comparison between the previous (i.e., exploratory) and current (i.e., confirmatory) samples when grouping participants transdiagnostically. No significant differences between groups or samples were observed. Bayes factor analyses also provided evidence supporting an absence of differences between samples in all cases: BFs between .10 (Guessing Condition, Patients) and .69 (Guessing Condition, HCs; Breath-hold Condition, Patients).

#### Interoceptive Precision

In Bayesian analyses predicting *IP* across the three HB tapping conditions in the current dataset (including group, condition, and their interaction as possible predictors), the transdiagnostic model (HCs vs. all patients) was favored over the diagnosis-specific model (BF = 43.23). Thus, we focus on transdiagnostic LMEs below (which allowed inclusion of the four individuals with anxiety alone). Analogous LMEs for diagnosis-specific groups, and associated Bayes factor analyses, are reported in **Supplementary Materials**. See Figure 4 and Figure 5 for plots of *IP* values by group and condition.

**Figure 5.**
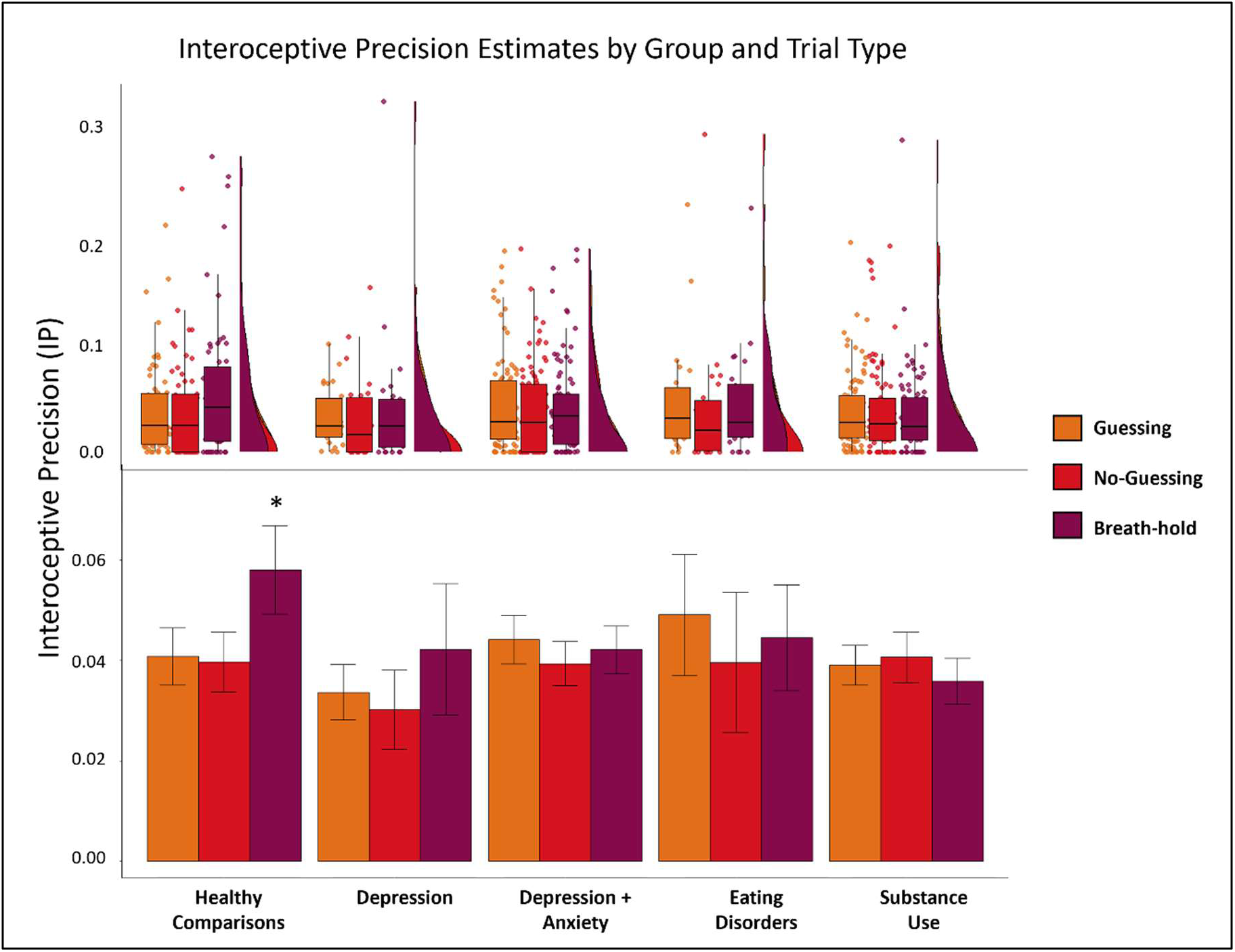
**Bottom**: Mean and standard error for interoceptive precision estimates by condition and clinical group. The black asterisk indicates that interoceptive precision (IP) was significantly greater in healthy comparisons than other groups (transdiagnostically) in the breath-hold condition, and healthy comparisons showed a significant increase in IP from the guessing and no guessing condition to the breath-hold condition that was absent in the other groups. **Top**: For more complete data characterization, we also show raincloud plots depicting the same results in terms of individual datapoints, boxplots (median and upper/lower quartiles), and probability densities.

The initial LME predicting *IP* (without covariates) revealed a marginal effect of condition (F(1,560) = 2.54, *p* = .080) and a marginal group by condition interaction (*F*(2,560) = 2.46, *p* = .086), but no main effect of group (*F*(1,280) = 2.03, *p* = .155). However, in the model accounting for planned covariates (age, sex, precision in the tone condition, BMI, median PTT, medication status, number of heartbeats, and interactions between number of heartbeats and group and condition separately), results showed a marginal effect of group (*F*(1,348) = 3.46, *p* = .064) and gained a significant group by condition interaction (*F*(2,569) = 3.11, *p* = .045). Post-hoc comparisons indicated that these results were explained by the following: 1) *IP* significantly increased in HCs in the breath-hold condition when compared to the two resting conditions (breath-hold: estimated marginal mean [EMM] = .058; guessing: EMM = .038; *t*(570) = 2.31, *p* = .021; no-guessing: EMM = .036, *t*(583) = 2.58, *p* = .010); and 2) *IP* during breath-hold was significantly greater in HCs than in the transdiagnostic patient sample (HCs: EMM = .058; patients: EMM = .040; *t*(806) = -2.56, *p* = .011). There were no significant differences between groups in the other conditions (*p* ≥ .698 in both cases). This LME also revealed a significant negative associations with number of heartbeats (*F*(1,348) = 6.90, *p* = .009) and precision in the tone condition (*F*(1,274) = 5.72, *p* = .017). All other effects were nonsignificant (*Fs* between .04 and 2.81, *ps* between .095 and .838).

#### Prior expectations

In Bayesian analyses predicting *pHB* across the three HB tapping conditions in the current dataset (including group, condition, and their interaction as predictors), a transdiagnostic model (HCs vs. all patients) had greater evidence when compared to a diagnosis-specific model (BF > 100). We therefore focus on transdiagnostic LMEs below (which allowed inclusion of the four individuals with anxiety alone). Analogous LMEs for diagnosis-specific groups are reported in **Supplementary Materials**. See Figure 4 and Figure 6 for plots of *pHB* values by group and condition.

**Figure 6.**
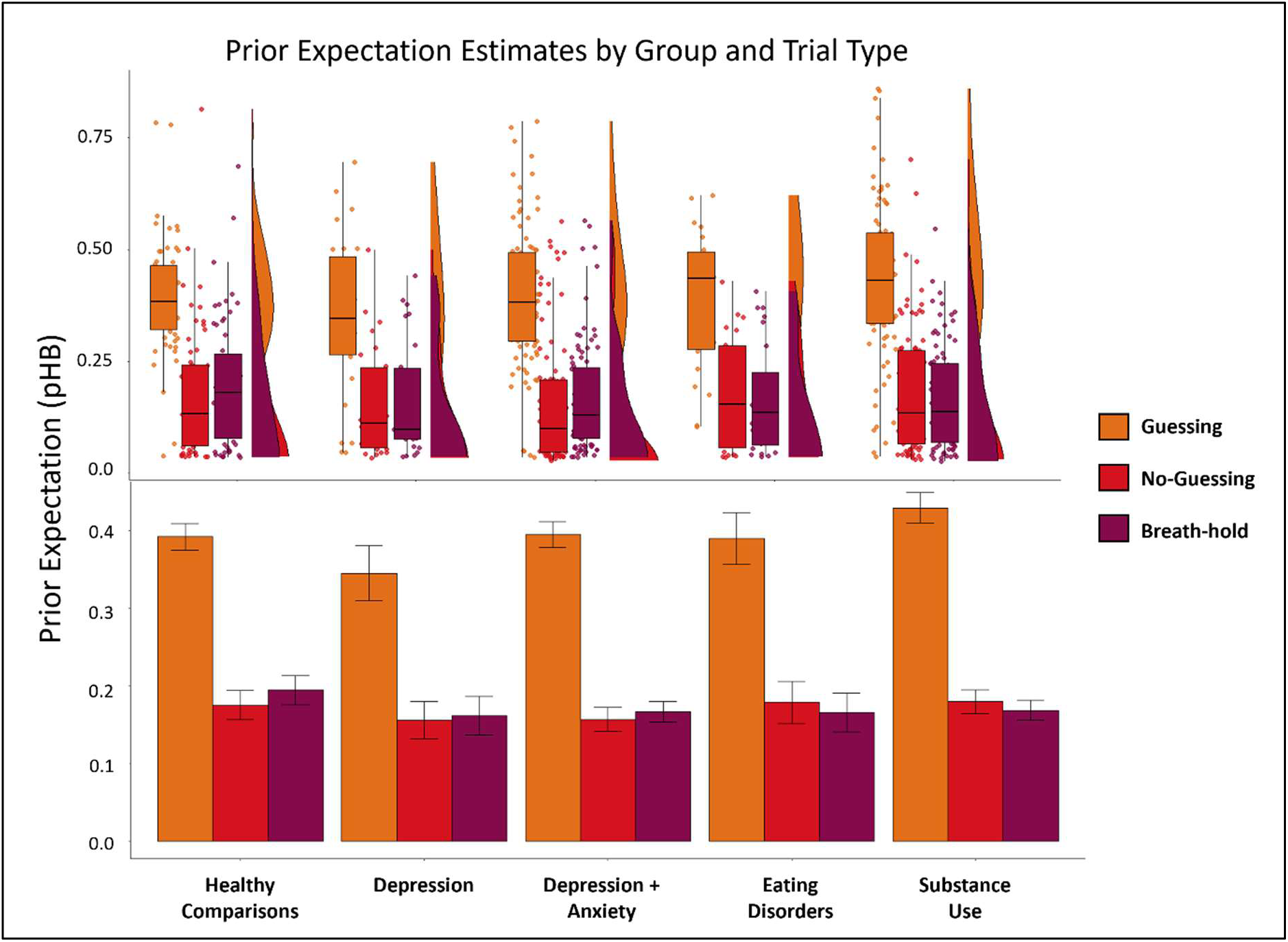
**Bottom**: Mean and standard error for *pHB* estimates by condition and clinical group. **Top**: For more complete data characterization, we also show raincloud plots depicting the same results in terms of individual datapoints, boxplots (median and upper/lower quartiles), and probability densities.

An initial LME predicting *pHB* (without covariates) revealed a main effect of condition (*F*(2,559) = 206.18, *p* < .001), but no main effect of group (*F*(1,281) = 0.33, *p* = .563) or a group by condition interaction (*F*(2,559) = 1.06, *p* = .349). Post-hoc comparisons indicated greater *pHB* in the guessing condition than in the other two conditions (guessing: EMM = .401; no-guessing: EMM = .176; *t*(519) = -17.44, *p* < .001; breath-hold: EMM = .182; *t*(530) = -16.88, *p* < .001). An analogous model including planned covariates confirmed the condition effect (*F*(2,596) = 20.74, *p* < .001) with the same pattern of results as previously mentioned. This model also revealed a marginally negative association with age (*F*(1,274) = 3.86, *p* = .050).There were no other significant effects (*F*s between 0.08 and 3.37, *p*s between .067 and .772).

#### Anticipating vs. reacting

All results for *AvR* are provided in **Supplementary Materials**. As in our prior study, there was no clear evidence for group differences. For plots of *AvR* by group and condition, see **Supplementary Figure S5**.

### Group by condition interactions in secondary measures

**Supplementary Figure S6** plots the values for all secondary measures in each condition within the previous and current datasets, separately for HCs and the combined patient group. As with model parameters, values for each measure were comparable. Bayes factor analyses provided evidence against the presence of differences between samples in all cases (BFs < 3), with the exception of self-reported confidence in the breath-hold condition for the transdiagnostic patient group, which was lower in the confirmatory sample (BF > 100).

In the current dataset, a Bayes factor analysis predicting heartbeat counting accuracy (including group, condition, and their interaction as predictors) found greater evidence for a model assuming transdiagnostic vs. diagnosis-specific effects (BF > 100).

An initial LME including only the effects of group, condition, and their interaction found evidence for a main effect of condition (*F*(2,562) = 19.34, *p* < .001) and a marginal effect of group (*F*(1,280) = 3.21, *p* = .074) on traditional heartbeat counting accuracy. Post-hoc contrasts revealed that accuracy scores were higher in the guessing condition than the two other HB conditions (guessing: EMM = .649; no-guessing: EMM = .274; *t*(559) = -5.15, *p* < .001; breath-hold: EMM = .336, *t*(560) = -4.29, *p* < .001). When accounting for covariates, there were also effects of group (*F*(1,334) = 6.68, *p* = .010) and condition (*F*(2,638) = 6.09, *p* = .002), but no interaction (*F*(2,568) = 0.52, *p* = .597). Post-hoc contrasts indicated that, again, accuracy was greater in the guessing condition than the other two conditions (guessing: EMM = .681; no-guessing: EMM = .316; *t*(559) = -5.32, *p* < .001; breath-hold: EMM = .367; *t*(567) = -4.60, *p* < .001) and that HCs had significantly greater accuracy than the transdiagnostic patient group across conditions (healthy: EMM = .524; patient: EMM = .385, *t*(273) = -2.31, *p* = .022). There was also a positive association with number of heartbeats (*F*(1,334) = 10.01, *p* = .002), a negative association with PTT (*F*(1,280) = 67.97, *p* < .001) and age (*F*(1,273) = 5.20, *p* = .023), and an effect of medication status (*F*(1,274) = 4.07, *p* = .045) such that those taking medication had better counting accuracy than participants who were not (*t*(274) = -2.02, *p* = .045). There were also significant interactions between number of heartbeats and condition (*F*(2,644) = 8.39, *p* < .001) and number of heartbeats and group (*F*(1,336) = 4.62, *p* = .032). Further post-hoc comparisons indicated that counting accuracy improved as the number of heartbeats increased in the guessing and no-guessing conditions compared to the breath-hold condition (breath-hold: EMM = -.002; guessing: EMM = .0130, *t*(667) = -3.74, *p* < .001; no-guessing: EMM = .011, *t*(641) = -3.25, *p* = .004) and that accuracy was higher in the patient groups as number of heartbeats increased (while this was not true for HCs; healthy: EMM = .002; patient: EMM = .012, *t*(336) = 2.15, *p* = .032).

Results of analogous LMEs with diagnosis-specific groupings are reported in the **Supplementary Materials**. To assess potential group differences in the effect of task condition on self-reported experience and physiology, we also carried out analogous LMEs assessing confidence, intensity, and difficulty, as well as heart rate. These results are reported in **Supplementary Materials**. No group by condition interactions in these variables were observed mirroring our *IP* results.

### Associations with symptom severity measures and interoceptive awareness scales

Given the heterogeneous nature of our clinical sample, we ran subsequent exploratory correlational analyses with continuous scores on some of the clinical measures gathered, excluding HCs, to assess whether model parameters might provide additional information about symptom severity in specific domains related to anxiety, depression, substance use, interoception, and emotional awareness. No significant relationships were found.

As in the previous paper, we examined correlations between model parameters and measures often thought to relate to interoception: the Multidimensional Assessment of Interoceptive Awareness (MAIA; (34)), the Toronto Alexithymia Scale (TAS-20; (39)), and the Anxiety Sensitivity Index (ASI; (36)). The results of these analyses are reported in **Supplementary Materials**.

### Combined sample analyses

After completing the pre-registered analyses above, we then combined the previous and current samples, which provided increased power to assess the possibility of narrower diagnostic differences. A specific diagnostic breakdown of the those with SUDs, and those with affective disorders without SUDs, in each sample is shown in Figure 6. In this combined sample, we tested if both model parameters (jointly) in each condition HB condition could predict the presence of specific substance use or affective disorders relative to HCs.

#### Substance use disorders

Comparison between specific disorders in iSUDs is shown in Figure 7. In models comparing specific SUDs to HCs (as shown in **Supplemental Table S2**) where sample size allowed, *IP* in the breath-hold condition could differentiate each SUD from HCs – including stimulant use disorder without comorbidities (Wald *z* = -4.18 to -2.42, *p* < .001 to .028). Prior beliefs (*pHB*) in the no-guessing condition differentiated those with cannabis use, opioid use, and sedative use (Wald *z* = 2.01 to 2.48, *p* = .013 to .044) from HCs, while in the breath-hold condition they could differentiate those with stimulant use, opioid use, and alcohol use (Wald *z* = 2.00 to 2.41, *p* = .016 to .046) from HCs. Still within iSUDs, *IP* in the breath-hold condition was also a significant differentiator for all affective disorders from HCs except for PTSD (Wald *z* = -3.52 to -2.16, *p* < .001 to .030). In the no-guessing condition, *pHB* uniquely differentiated generalized anxiety disorder (GAD) from HCs (Wald *z* = 2.75, *p* = .006). In all significant cases, iSUDs with specific diagnoses had lower interoceptive precision and higher prior bias estimates than HCs. When comparing each specific disorder to others in the substance use group (**Supplemental Table S3**), model parameters were unable to differentiate any disorders, with the exception of *pHB* in the no-guessing condition, which was able to differentiate GAD from other disorders (Wald *z* = 2.21, *p* = .027) due to significantly higher prior bias estimates in those with GAD.

**Figure 7.**
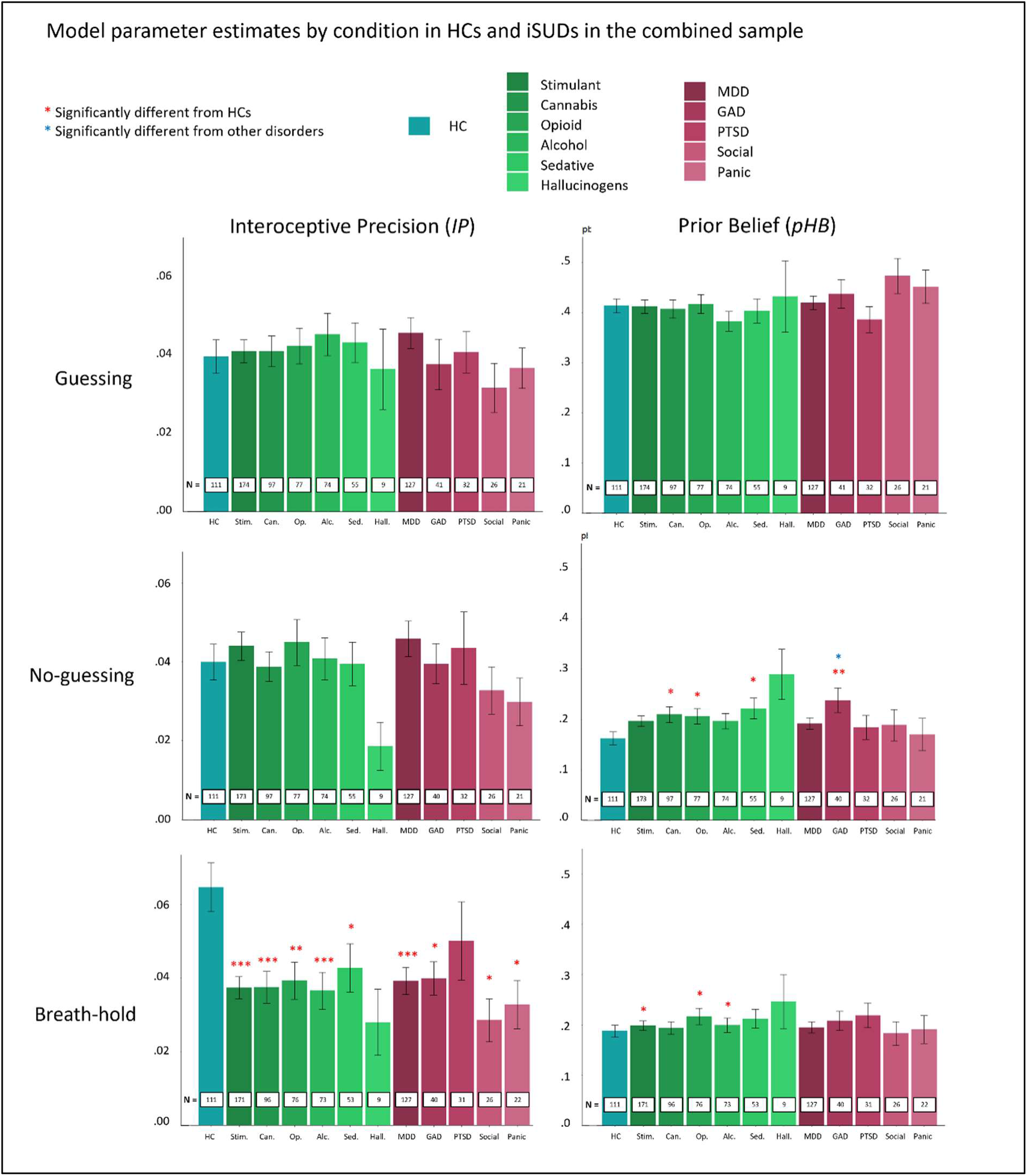
Model parameter estimates (*IP* and *pHB*) in the combined exploratory/confirmatory dataset in the three HB tapping conditions in HCs and iSUDs, separated by specific diagnostic groupings. Note that these groupings contain overlapping participants as they include anyone with a given diagnosis irrespective of co-morbid diagnoses. Red asterisks indicate groups differentiable from HCs in logistic regressions. Blue asterisks indicate diagnostic groups that could be differentiated from other diagnoses in similar logistic regressions (excluding HCs). Detailed results of these analyses can be found in **Supplementary Tables S2** and **S3**.

#### Affective disorders without SUDs

Comparison between specific disorders in individuals with affective disorders without SUDs (iDEP, iANX, and iDEP+ANX; iADs) is shown in Figure 8. As detailed in **Supplemental Tables S4** and **S5**, we found that *IP* estimates in the breath-hold condition differentiated HCs from most diagnoses: MDD, GAD, panic, and PTSD (Wald *z* = -3.78 to -2.38, *p* < .001 to .017) where iADs had lower precision estimates than HCs. In the no-guessing condition, *IP* also differentiated those with panic disorder from HCs (Wald *z* = -2.02, *p* = .044), again, due to significantly lower estimates in those with panic disorder. When comparing affective disorders to each other (excluding HCs), we found that 1) *IP* in the no-guessing condition differentiated GAD from other disorders (Wald *z* = 2.31, *p* = .021), and 2) *IP* in the breath-hold condition differentiated social anxiety from other disorders (Wald *z* = 2.18, *p* = .029). In both cases, precision estimates were higher than other disorders (i.e., closer to HCs). Prior bias, *pHB*, did not differentiate any specific AD diagnoses from HCs.

**Figure 8.**
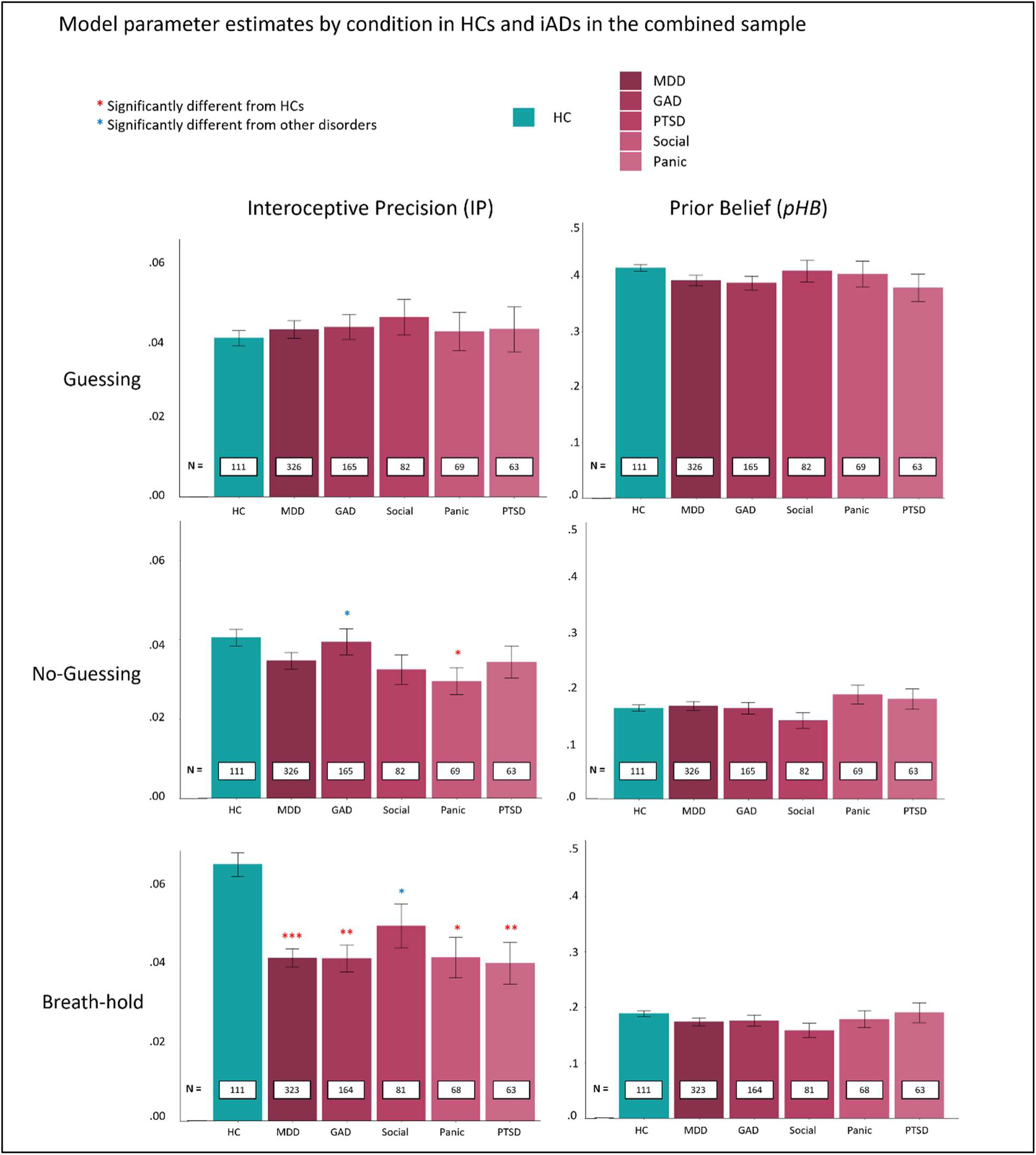
Model parameter estimates (*IP* and *pHB*) in the three HB tapping conditions in HCs and iADs separated by specific diagnostic groupings.

#### Eating disorders

When comparing participants with specific EDs (anorexia nervosa vs. bulimia nervosa) to HCs, and to each other (**Supplemental Tables S6** and **S7**), parameter estimates did not differentiate groups (while noting the small sample sizes in these specific groups).

#### Predictive Categorization

Because of the transdiagnostic focus of our previous results, we first tested the predictive categorization ability of a model (with *IP* parameters in all three HB conditions as predictors) that was trained on the exploratory sample to see how well it could classify HCs vs. all patients together (as shown in Figure 9). This model had a predictive accuracy of .65 with an AUC of .57, indicating low discriminability with *IP*. Results of testing predictive models on the classification of each patient group (excluding iANX) separately are also shown in Figure 9. Accuracies were between .51 and .62 indicating, again, low discriminability with *IP*. However, all AUCs remained above chance (.55 - .58).

**Figure 9.**
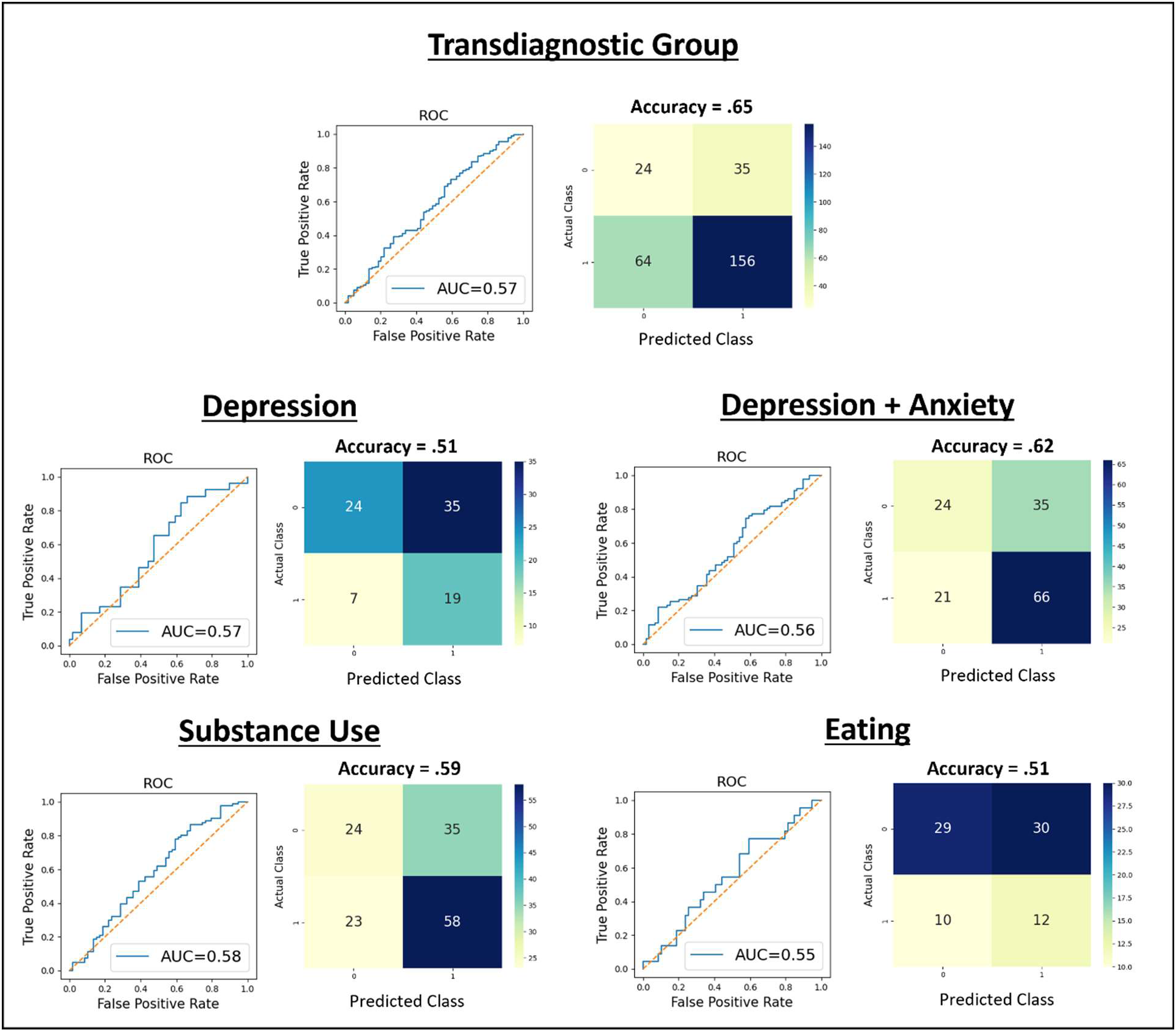
Results of logistic regression models trained on the exploratory sample and then tested on the confirmatory sample (classifying individuals as HCs vs. each patient group). Receiver operating characteristic curves (ROCs) illustrate the true positive rate (sensitivity) vs. false positive rate (1 – specificity) for different categorization thresholds. Performance is quantified by associated area-under-the-curve (AUC) scores, reflecting how often a random sample will be assigned to the correct group with higher probability. Acceptable AUCs vary by application, but the following heuristic cutoff values have been proposed: 0.5 = No discrimination, 0.5-0.7 = Low discrimination, 0.7-08 = Acceptable discrimination, 0.8-0.9 = excellent discrimination, > 0.9 = Oustanding discrimination (40). Accuracy at a neutral threshold of 0.5 is also shown in a dotted orange line.

## Discussion

### Replication of Primary Findings

In this report, we replicated the computational modeling results of our prior study (15) examining behavior on a heartbeat tapping task in a transdiagnostic psychiatric patient sample, including individuals with anxiety, depression, substance use, and/or eating disorders. Most centrally, we sought to replicate the finding that, across clinical groups, patients did not assign greater precision (*IP*) to interoceptive signals during an inspiratory breath-hold manipulation designed to increase the magnitude of these signals. This contrasted with healthy participants, who did show these increases – suggesting greater interoceptive awareness when signals change (also observed in a second study; (16)). As expected, the healthy and patient groups in this new sample showed this same pattern. Thus, the group differences found in our previous report successfully replicated.

Bayesian analyses provided evidence against specific clinical group differences and instead favored a model assuming the difference in *IP* was transdiagnostic. As also observed previously, while prior biases (*pHB*) were sensitive to task instructions, there was no evidence of differences in these biases (or effects of instructions) between healthy comparisons and the transdiagnostic patient sample. This further confirmed that the observed interoceptive abnormality was due to blunted signal precision estimates and not biased (or rigid) prior beliefs.

### Replication of Secondary Findings

As further validation of the model parameters, *IP* estimates again showed positive relationships with self-report ratings of heartbeat intensity, while *pHB* was lower in those who reported greater task difficulty. Prior bias estimates also showed strong associations with counting accuracy, consistent with previous concerns that the latter metric reflects beliefs about heart rate as opposed to detection accuracy (9, 14, 41). Notably, unlike in our previous report, we did also observe greater counting accuracy in HCs within the confirmatory sample.

When combining samples, which maximized the power to detect potential differences between narrower diagnostic categories, we also found a few notable results. Namely, logistic regressions within iSUDs found that *IP* in the breath-hold condition differentiated HCs from each substance use and affective disorder (with the exception of PTSD); but did not differentiate between disorders. This suggested the same transdiagnostic interpretation of *IP* results described above. Unlike the broader results above, these regressions found that *pHB* in the no-guessing condition also differentiated HCs from some disorders in iSUDs: cannabis use disorder, opioid use disorder, sedative use disorder, and GAD (i.e., greater values in each disorder than HCs). These findings may be consistent with previous models of drug craving in iSUDs that suggest over-weighting of prior expectations regarding physiological states and subsequent down-weighting of interoceptive signals (42, 43). These *pHB* estimates also distinguished iSUDs with GAD from those with other disorders (higher values in GAD), suggesting some potential diagnostic specificity not detected previously.

In affective disorders without SUDs (iADs), *IP* – again, in the breath-hold condition – differentiated each affective disorder from HCs except social anxiety; it also differentiated those with social anxiety from all other affective disorders. As these findings highlight social anxiety in particular, future research should test if precision-weighting in social anxiety is reliably comparable to HCs. Unlike in iSUDs, *pHB* did not differentiate any disorders from HCs or from each other.

Finally, after training a model on the exploratory sample and testing its predictive accuracy when classifying participants by diagnostic status in the confirmatory sample, we found above chance, but generally low discriminatory power, with the best predictive accuracy in differentiating patients transdiagnostically. Thus, while potentially insightful regarding mechanistic differences in subpopulations within HCs and patient groups, these findings may be of limited predictive clinical utility.

### Comparative Insights and Clinical Implications of Replicated Findings

The present replication findings align with a recent report (44) that examined cardiac interoception (via heartbeat counting and heartbeat detection tasks) in individuals with various psychiatric conditions including affective disorders, personality disorders, and psychosis spectrum disorders. Both studies affirm the transdiagnostic importance of cardiac interoception to psychopathology and find abnormalities in interoceptive processing when compared to healthy or non-clinical participants. Additionally, both studies highlight the impact of these interoceptive deficits on affective symptoms like anxiety and depression. However, several unique aspects distinguish the current study. First, we utilized an interoceptive perturbation technique, involving breath-holding, to reveal specific abnormalities in interoceptive processing. Second, the Bayesian computational model allowed for a nuanced understanding of deficits in the precision-weighting of afferent cardiac signals. Third, our study benefits from a larger overall sample size. Lastly, the pre-registered approach confirms and extends our original findings, suggesting that these deficits are likely to be consistently observed across varying psychiatric samples, such as those with mood/anxiety, substance use, and eating disorders. Collectively, these studies contribute to a growing body of evidence supporting the role of abnormal interoceptive processing in psychiatric disorders, as also demonstrated by recent studies in generalized anxiety disorder (45, 46) and eating disorders (47–49). Given these converging findings, there is increased impetus for the translation of this knowledge into clinical practice. Future work might explore the utility of Bayesian modeling as a tool for longitudinally assessing interoceptive dysfunctions across various psychiatric disorders. Furthermore, interoceptive training interventions could be developed to target precision-weighting of cardiac signals, potentially offering a novel therapeutic approach for enhancing visceral regulation in affected individuals (see (50–54) for examples of some candidate approaches impacting the cardiac domain). These interventions may be especially pertinent for patient populations demonstrating significant interoceptive deficits, such as those with anxiety, substance use, and eating disorders, and could provide an initial framework for developing targeted therapeutic strategies.

### Strengths, Limitations, and Conclusion

This study had most of the same strengths and limitations as our prior report. Strengths included the large sample size, the novel computational modeling approach, and the use of specific task conditions to selectively probe changes in distinct computational mechanisms and address concerns associated with other interoception measurement approaches. The task itself is also less time-consuming for participants than other common tasks, with each task condition lasting only 60 seconds. However, it is also possible that longer testing durations (e.g., a greater number of 1-minute trials per condition) could improve parameter estimates during perceptual modelling. Another limitation is that, because our modelling approach required distinct heartbeat states for comparison to behavior, we had to make choices about perceptual windows based on EKG signals. As mentioned in our prior report, other choices for defining these windows are possible.

For replication purposes, we used the same model as in our prior report; however, additional free parameters and model structures could be considered in future work. For example, alternative models (e.g., the Hierarchical Gaussian Filter; HGF) that also capture internal perceptual uncertainty could be considered (55). Lastly, the small sample sizes of the anxiety and eating disorder groups limited their involvement in group-comparison analyses and the confidence with which the results can be interpreted. Future work could focus on these anxiety and eating disorders to test if the transdiagnostic effects suggested here generalize to these groups.

## Conclusion

In summary, this study replicated several effects found in a prior study comparing healthy individuals to a transdiagnostic patient sample using computational modeling of behavior on a heartbeat perception task. These results confirm a transdiagnostic failure to adapt beliefs about the precision of interoceptive signals, which could represent a vulnerability and/or maintenance factor for psychopathology broadly. Future research should investigate underlying neural and peripheral physiological mechanisms and evaluate whether this newly identified difference may be effectively targeted by current or novel clinical interventions.

## Data Availability

All data produced in the present study are available upon reasonable request to the authors.

## Software Note

All model simulations were implemented using standard routines (**spm_MDP_VB_X.m**) that are available as Matlab code in the latest version of SPM academic software: http://www.fil.ion.ucl.ac.uk/spm/. The specific code used for our model can be found in supplementary materials.

## Acknowledgements

Other contributors to the Tulsa 1000 project include the following: Robin L. Aupperle, Justin S. Feinstein, Jonathan B. Savitz, and Teresa A. Victor. Teresa A. Victor is the lead author for this group and can be reached at.

## Supplementary Files

**Supplementary materials.** This file includes results of supplementary analyses as well as supplementary figures, as referred to in the main text.

**Supplementary code.** This file contains the MATLAB code used for modelling task behavior.

**Study data.** This file includes all data used in the analyses reported in the manuscript.

## Supplemental Materials

### Supplementary Figures

**Figure S1.**
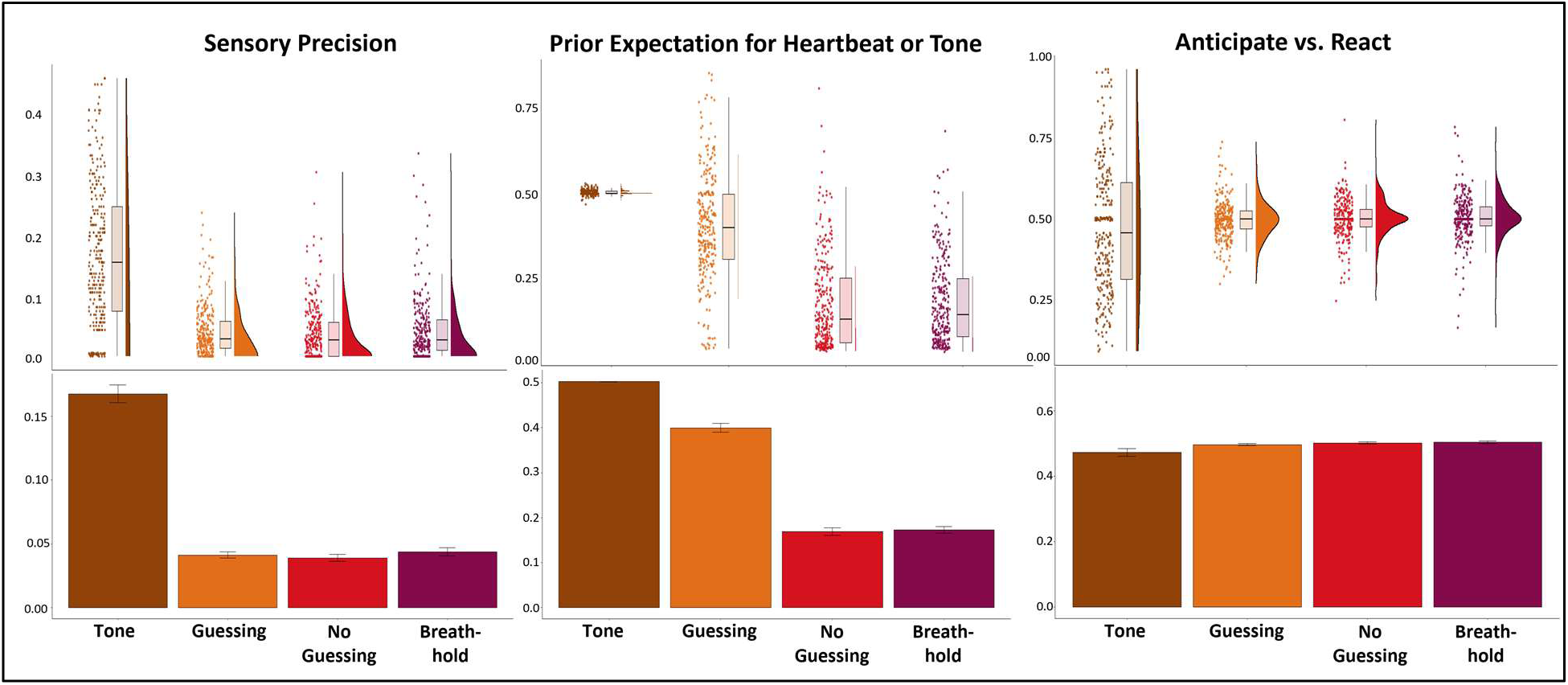
**Bottom:** Bar plots comparing means and standard errors for model parameters in the tone condition to those in the HB conditions. Prior beliefs for tones reflected no bias. Sensory precision for the tone condition was high, which was expected given the unambiguous nature of this signal relative. The Anticipate vs. React parameter in the tone condition did not significantly differ from the HB conditions, but had greater variance. **Top**: For more complete data characterization, we also show raincloud plots depicting the same results in terms of individual datapoints, boxplots (median and upper/lower quartiles), and distributions. These further illustrate that, for the Anticipate vs. React parameter, nearly equally sized clusters of participants appeared to adopt more anticipatory (<.5) vs. reactive (>.5) strategies in the tone condition, and that prior beliefs remained unbiased (.5; with little variance) in the tone condition relative to the HB conditions. This pattern of results matched that found in our previous study.

**Figure S2.**
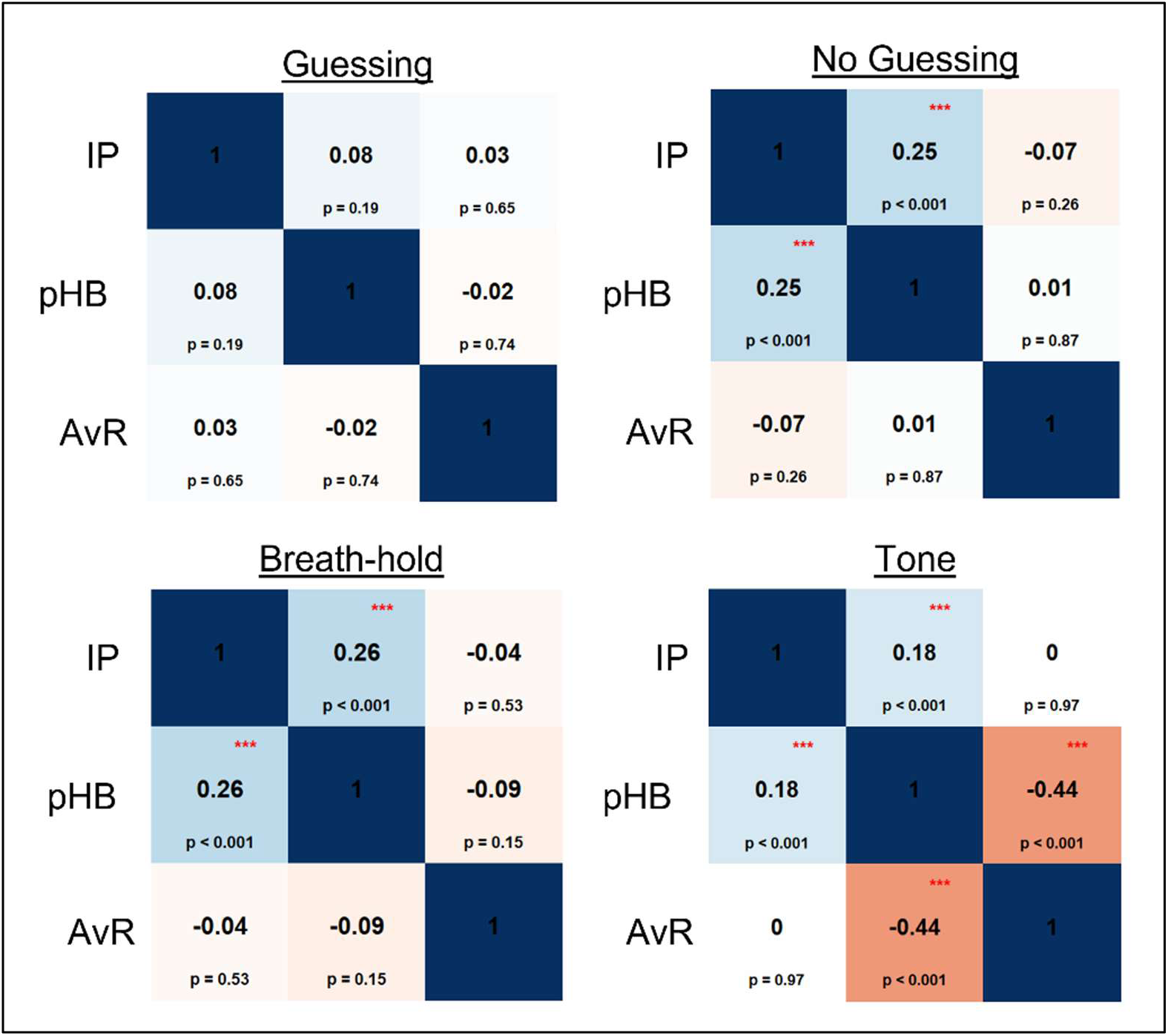
Correlations between model parameters across conditions. *IP* = interoceptive precision (or sensory precision in the tone condition), *pHB* = prior expectation over heartbeats (or tone), *AvR* = anticipating vs reacting.

**Figure S3.**
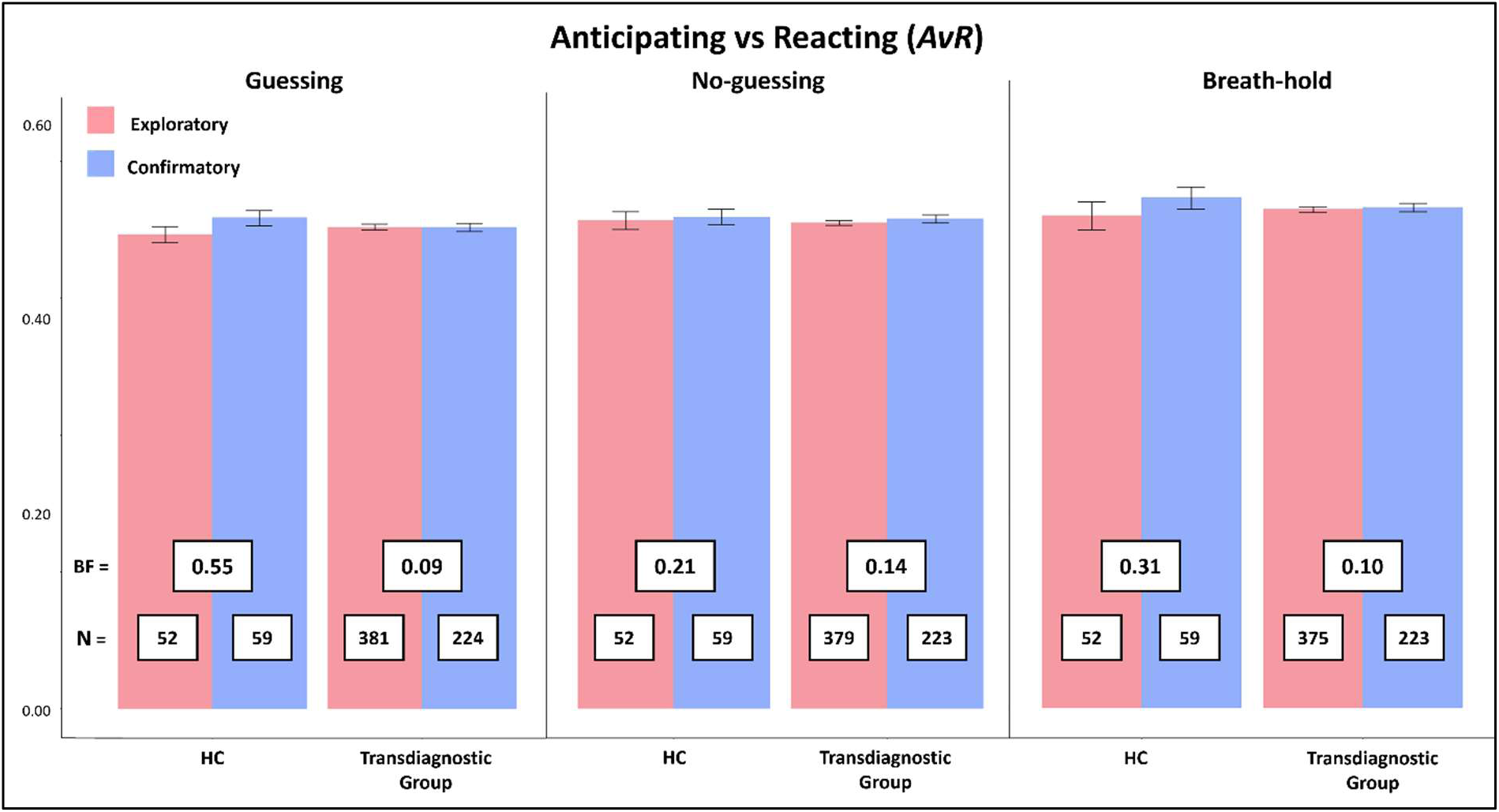
Mean and standard error for Anticipating vs Reacting (*AvR*) in the three heartbeat tapping conditions. Bayes factors (BFs) comparing samples by group and condition revealed evidence favoring models without a difference between samples in all cases. Linear models (LMs) predicting *AvR* in each condition with group and sample found no significant effects or interactions (*F* ≤ 1.95, *p* ≥ .163 in all cases).

**Figure S4.**
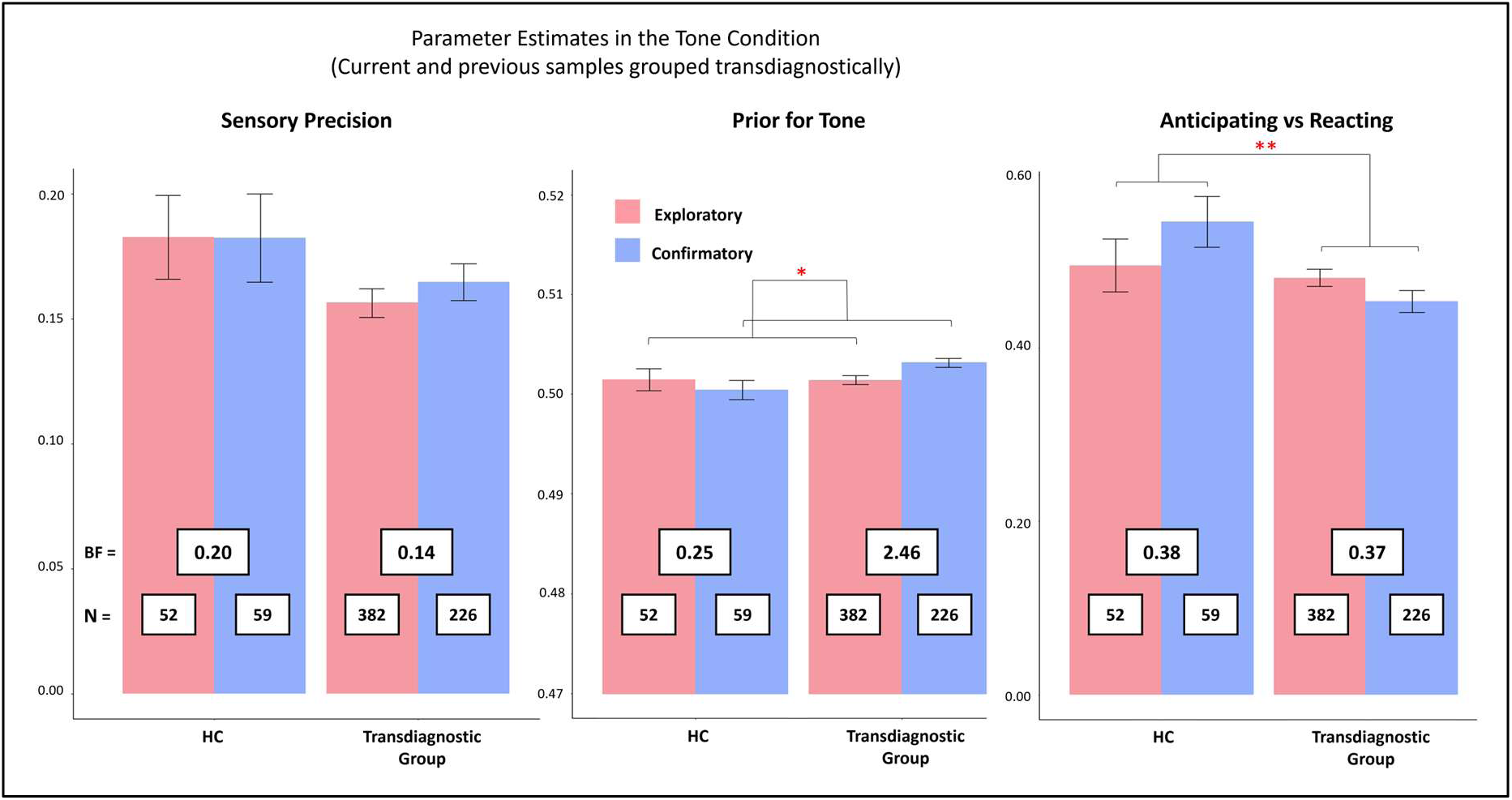
Mean and standard error for model parameters by group and sample in the tone condition. Bayes factors (BFs) comparing samples by group and condition revealed evidence favoring models without a difference between samples in all cases. Linear models (LMs) predicting each parameter in the tone condition with group and sample found a significant effect of sample for *pTone* (*F*(1,715) = 4.52, *p* = .034) such that estimates in the confirmatory sample were higher than the exploratory sample, and a significant effect of group for *AvR* (*F*(1,715) = 6.82, *p* = .009) indicating that healthy controls (HCs) were more reactionary than patients. No significant effects were found for sensory precision (*F*s ≤ 3.36, *p*s > .067).

**Figure S5.**
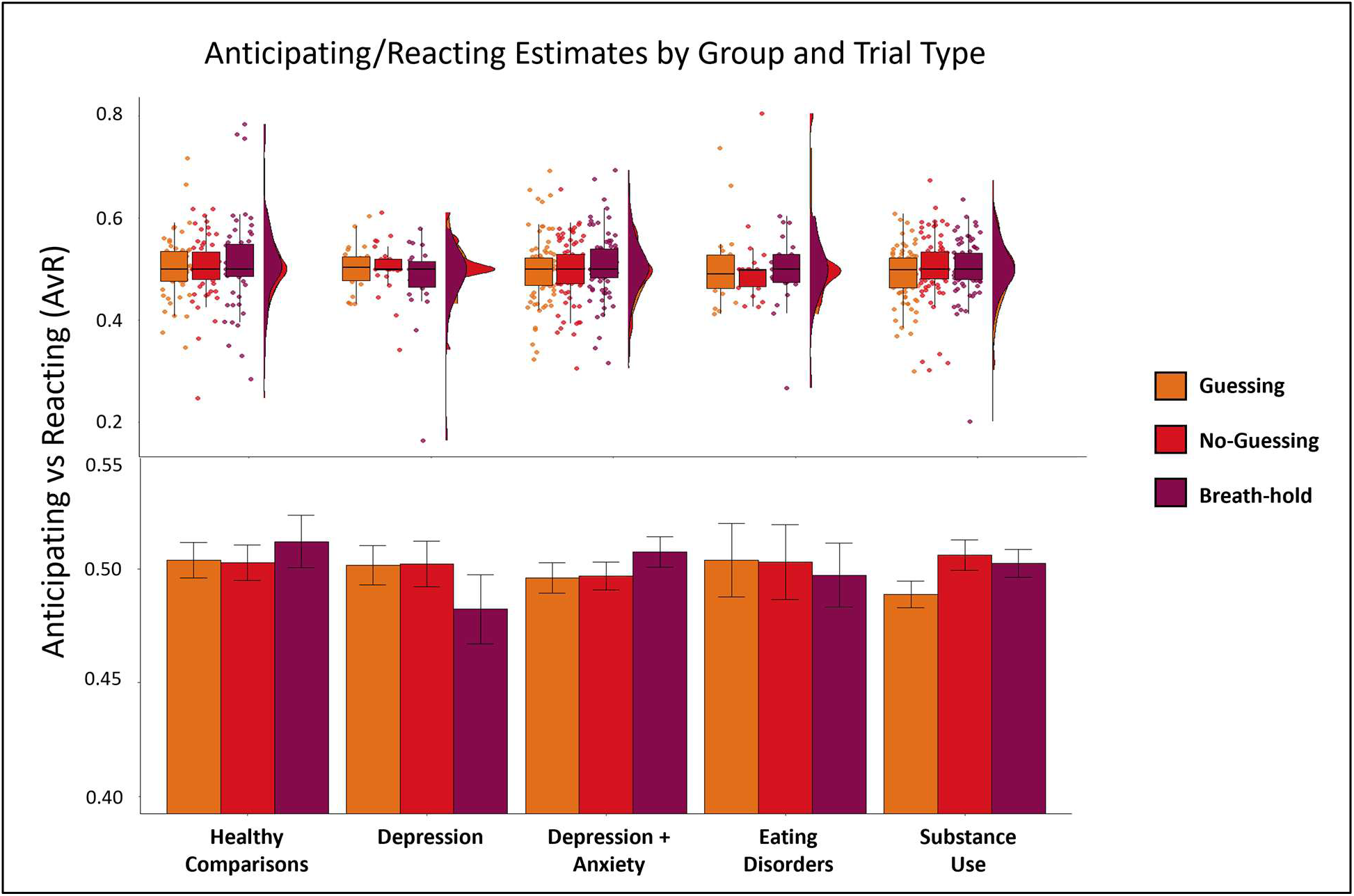
**Bottom**: Mean and standard error for Anticipating vs Reacting (*AvR)* estimates by condition and clinical group. **Top**: For more complete data characterization, we also show raincloud plots depicting the same results in terms of individual datapoints, boxplots (median and upper/lower quartiles), and probability densities.

**Figure S6.**
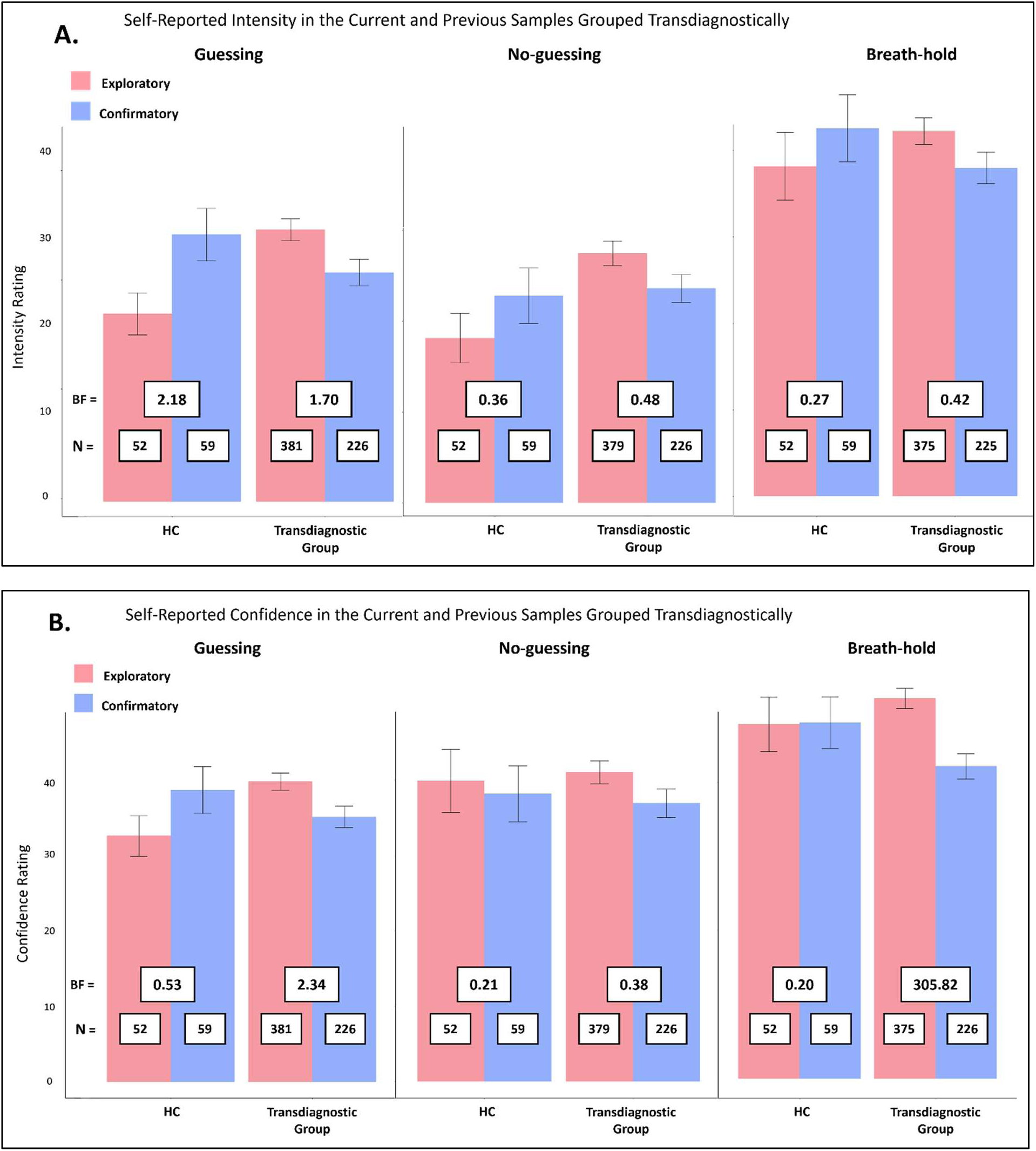
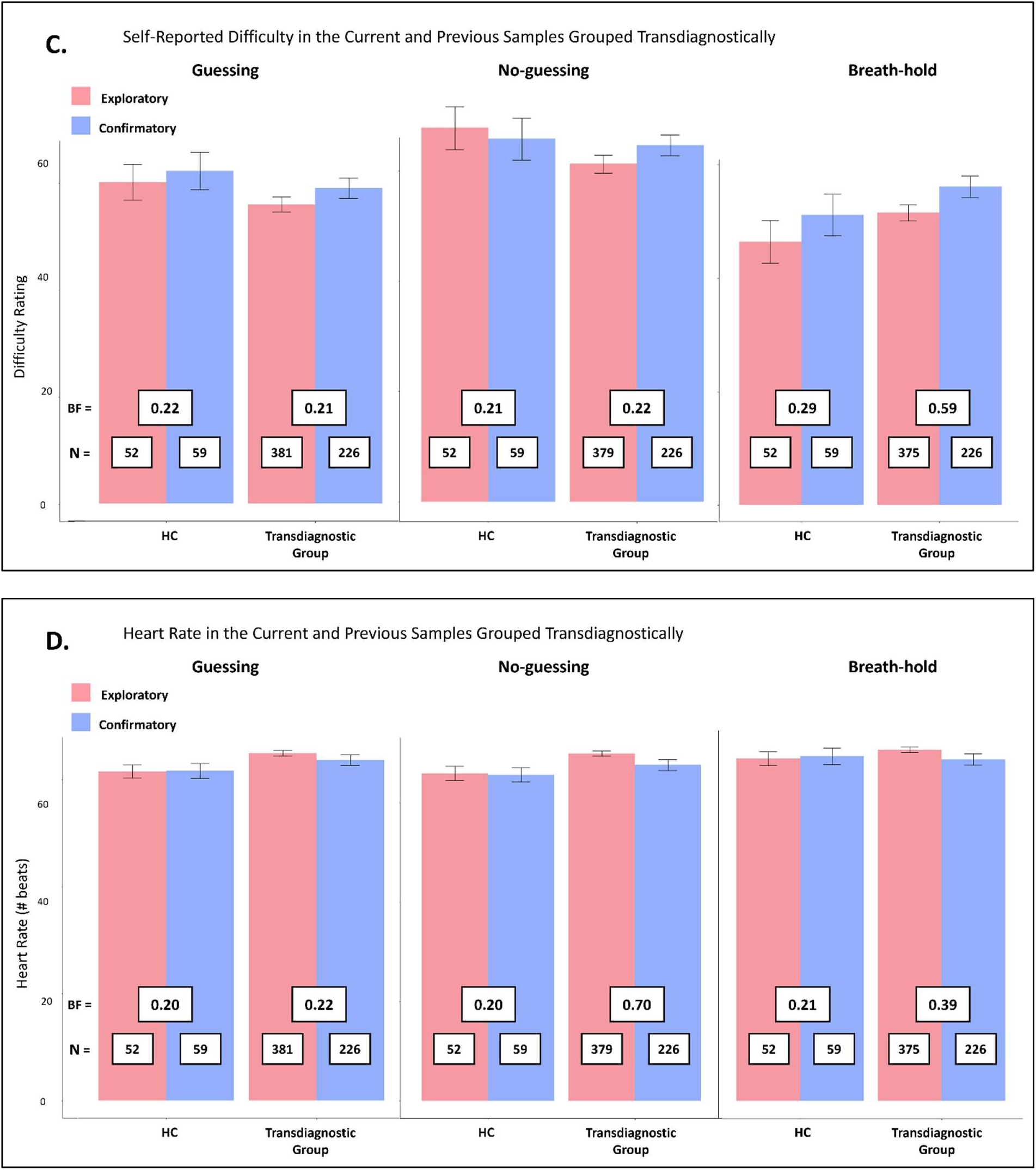
Self-reported task ratings in the confirmatory and exploratory samples across the three heartbeat tapping conditions separated into healthy controls (HCs) and the transdiagnostic patient group. Bayes factors (BFs) comparing samples by group and condition revealed evidence favoring models without a difference between samples in most cases. However, some evidence was found suggesting lower confidence ratings in the patient group within the confirmatory sample in the guessing and breath-hold conditions, and for different intensity ratings in both groups within the guessing condition (greater in HCs and weaker in patients in the confirmatory sample).

### Supplementary Analyses

#### Interoceptive Precision

In an initial LME predicting *IP* using diagnosis-specific groupings, now excluding the 4 iANX, there were no significant group or condition effects (*F*s < 1.11, *p*s > .331). Accounting for age, sex, precision in the tone condition, BMI, median PTT, medication status, number of heartbeats, and interactions between number of heartbeats and group and condition separately, there were negative associations with tone precision (*F*(1,265) = 5.20, *p* = .023) and number of heartbeats (*F*(1,367) = 4.66, *p* = .031).

In transdiagnostic JZS Bayes Factor comparisons including all models including group, condition, and/or ID predicting *IP*, the winning model contained only group (where HCs had higher values) but with limited evidence relative to a model that only contained ID (BF = 2.40). When including covariates, the winning model included only tone precision and ID (where tone precision was positively associated with *IP* estimates), but the data provided less evidence for this model than a null model containing only ID (BF = .62). Analogous comparisons separating participants by specific diagnoses but removing covariates found that the winning model contained condition only and was not favored over the model with only ID (BF = .25). Finally, including covariates led to a winning model identical to that for the transdiagnostic grouping; the best model contained age, tone precision, and ID, yet the ID-only model was still favored (BF = .36).

#### Prior Expectations

In an LME with diagnosis-specific groups without covariates, there was a significant effect of condition (*F*(2,549) = 220.14, *p* < .001), but not group (*F*(4,277) = 0.93, *p* = .445), or a group by condition interaction (*F*(8,548) = 0.90, *p* = .519). Accounting for differences in age, sex, precision in the tone condition, BMI, median PTT, medication status, number of heartbeats, and interactions between number of heartbeats and group and condition separately confirmed the condition effect (*F*(2,582) = 19.85, *p* < .001) and revealed a marginal negative association with age (*F*(1,264) = 3.42, *p* = .065) and number of heartbeats (*F*(1,437) = 3.85, *p* = .050). In both models, post-hoc comparisons revealed that the condition effect was, again, driven by significantly greater *pHB* values in the guessing condition than in the other two conditions (*p*s < .001).

Bayes Factor comparisons of transdiagnostic models predicting *pHB* that contained combinations of group, condition, and/or ID found that the winning model contained condition (where estimates were higher in the guessing condition than the other two conditions) and ID and was preferred over a model containing only ID (BF > 100). When including covariates, the winning model contained condition (indicating that, again, estimates were higher in the guessing condition than the other two conditions) and ID as well as a group by heart rate interaction (reflecting the fact that *pHB* estimates were negatively associated with heart rate in patients but not in HCs) and was favored over the model containing only ID (BF > 100). Subsequently separating participants into diagnosis-specific groupings and removing covariates found that the best model again contained condition (with higher estimates in the guessing condition) and ID – as with the transdiagnostic grouping – and evidence strongly favored this model over the model containing only ID (BF > 100). After including covariates, evidence best supported a model that included condition (where estimates were higher in the guessing condition), number of heartbeats (where *pHB* was negative associated with heart rate), and ID and was favored over the model that included only ID (BF > 100).

#### Anticipating vs Reacting

In Bayesian analyses predicting *AvR* across the three HB tapping conditions (including group, condition, and their interaction), the models that grouped participants transdiagnostically and by diagnostic groups were given equivalent evidence (BF = 1.29). Thus, for simplicity, we focus first on the transdiagnostic LMEs as with the other parameters, but include diagnosis-specific models for completeness.

An initial LME that grouped participants transdiagnostically (and included iANX), there were no significant effects of group, condition, or group by condition interaction (*F*s < 1.53, *p*s > .218 for each). Including covariates, there were, again, no significant effects of any predictor (*F*s < 1.79, *p*s > .182 in all cases). Subsequently, in an LME including diagnosis-specific groupings and no covariates, there were no significant effects (*F*s < 0.82, *p*s > .586). However, an analogous LME including covariates found an effect of group (*F*(4,373) = 2.54, *p* = .039) and a group by heart rate interaction (*F*(4,378) = 2.55, *p* = .039), though post-hoc contrasts indicated no significant differences between groups (*p*s ≥ .336) and only a marginal difference between iDEP+ANX and iSUDs by heart rate (*t*(417) = 2.71, *p* = .054) such that iSUDs reacted more as heart rate increased while iDEP+ANX reacted less.

Finally, in Bayes Factor comparisons of models including combinations of group, condition, and ID predicting *AvR*, we found that the winning model in the transdiagnostic grouping included an effect of group (where HCs had higher estimates than patients) and was favored over the model containing just ID (BF = 2.16). After including covariates, the most evidence was afforded to a model containing only sex, but the model containing ID alone was favored (BF = .27). An analogous comparison of models without covariates while grouping participants by diagnosis found the best evidence for a model including only condition but this model was not favored over a model containing only ID (BF = .266). Including covariates led to a winning model containing only age but was not favored over the model containing ID alone (BF = .22).

#### Other task measures

After performing LMEs predicting the traditional counting accuracy metric with the transdiagnostic grouping, we then separated the patient groups by diagnosis. In an initial LME without covariates, there was a main effect of condition (*F*(2,550) = 13.72, *p* < .001), such that accuracy was greater in the guessing condition (EMM = .621) than the two other conditions (no-guessing: EMM = .282, *t*(545) = -4.81, *p* < .001; breath-hold: EMM = .325, *t*(552) = -4.22, *p* < .001). When including covariates, there was a main effect of group (*F*(4,352) = 11.21, *p* < .001) and condition (*F*(2,621) = 6.24, *p* = .002), but no group by condition interaction (*F*(8,550) = 0.95, *p* = .473). Post-hoc comparisons found that iDEP+ANX (EMM = .328) were significantly different from HCs (EMM = .505, *t*(263) = 2.65, *p* = .009). No other group differences were present. There were also significant differences between the guessing condition (EMM = .629) and the two other conditions such that accuracy was highest in the guessing condition (no-guessing: EMM = .300, *t*(544) = 5.08, *p* < .001; breath-hold: EMM = .342, *t*(551) = 4.46, *p* < .001). There were additional negative associations with PTT (*F*(1,270) = 69.77, *p* < .001) and age (*F*(1,263) =, *p* = .032), as well as interactions between number of heartbeats and condition (*F*(2,625) = 8.29, *p* < .001) and number of heartbeats and group (*F*(4,356) = 10.22, *p* < .001). Post-hoc comparisons showed that this interaction was driven by a negative relationship between number of heartbeats in the breath-hold condition and counting accuracy (breath-hold: ET = - .004) and positive relationships in the guessing (ET = .01, *t*(648) = 3.63, *p* < .001) and no-guessing conditions (ET = .008, *t*(622) = 3.35, *p* = .003). However, when separating the patient groups, these comparisons indicated that the number of heartbeats by group interaction was driven by significant differences in iDEP+ANX (EMM = .023) and iDEP (EMM = -.004, *t*(385) = -4.02, *p* < .001), iSUD (EMM = .007, *t*(388) = 4.61, *p* < .001), iED (EMM = -.006, *t*(354) = 3.78, *p* = .002), and HC (EMM = .002, *t*(352) = 4.23, *p* < .001).

In analogous LMEs, we investigated potential group differences in the self-report measures of heartbeat intensity, confidence, and difficulty, as well as heartrate. These models were first examined only including condition, group, and their interaction as predictors and then were performed again including the same covariates as in our previous analyses. These models, as before, excluded the tone condition.

##### Self-reported heartbeat intensity

Bayesian analyses strongly favored the transdiagnostic model over the diagnosis-specific model (BF > 100). Grouping participants transdiagnostically (HCs vs. all patients, including iANX), an initial LME predicting self-reported intensity indicated significant effects of condition (*F*(2,565) = 45.04, *p* < .001). Post-hoc contrasts revealed that each condition was different from the others (*p* < .03 in all cases) such that intensity ratings were highest in the breath-hold condition and lowest in the no-guessing condition. A subsequent LME containing covariates again revealed a significant effect of condition (*F*(2,601) = 9.13, *p* < .001), as well as a positive effect of number of heartbeats (*F*(1,488) = 7.09, *p* = .008) and a marginal effect of sex(*F*(1,276) = 3.58, *p* = .060). Post-hoc contrasts confirmed that intensity was greater in the breath-hold condition (EMM = 41.1) than in the guessing condition (EMM = 28.8, *t*(583) = 6.51, *p* < .001) which was greater than intensity ratings in the no-guessing condition (EMM = 25.0; *t*(566) = 2.04, *p* = .042) and that male participants gave marginally higher intensity ratings (EMM = 34.1) than female participants (EMM = 29.1, *t*(276) = 1.89, *p* = .060).Separating groups by diagnosis (removing the 4 iANX), we found further evidence for a main effect of condition (*F*(2,552) = 39.28, *p* < .001) such that intensity ratings in the breath-hold condition (EMM = 38.2) were significantly higher than those in the other two conditions (guessing: EMM = 25.2; *t*(552) = 7.32, *p* < .001; no-guessing: EMM = 23.9, *t*(552) = 8.01, *p* < .001). When including covariates, there was an effect of condition (*F*(2,585) = 7.29, *p* < .001) and a marginal positive association with number of heartbeats (*F*(1,577) = 3.75, *p* = .053). Post-hoc contrasts revealed that these results were driven by higher intensity ratings in the breath-hold condition than the other two conditions (*p* < .001 in both cases).

##### Self-reported confidence

Bayes factor analyses found greater evidence for a model that grouped participants transdiagnostically than by separate diagnosis-specific groups (BF > 100). Grouping participants transdiagnostically, we found a significant effect of condition (*F*(2,566) = 8.84, *p* < .001), such that confidence ratings in the breath-hold condition (EMM = 45.1) were significantly higher than the other two HB conditions (guessing: EMM = 37.0, *t*(566) = 3.78, *p* < .001; no-guessing: EMM = 37.6; *t*(566) = 3.49, *p* < .001). In a subsequent LME that included covariates, there was a marginal effect of condition (*F*(2,612) = 2.85, *p* = .059) and an effect of number of heartbeats (*F*(1,440) = 5.88, *p* = .016). Confidence ratings were greater in the breath-hold condition (EMM = 45.2) than the other two conditions (guessing: EMM = 37.7; *t*(581) = 3.45, *p* < .001; no-guessing: EMM = 38.6; *t*(598) = 3.03, *p* = .003) and were associated with a greater number of heartbeats. When separating the patient groups by diagnosis and removing iANX, there were main effects of condition (*F*(2,552) = 5.23, *p* = .006) and group (*F*(4,276) = 3.36, *p* = .010). Post-hoc contrasts revealed that, again, ratings were higher in the breath-hold condition than the others, that iDEP (EMM = 32.8) and iED (EMM = 28.6) were less confident than iSUD (EMM = 42.1) and HCs (EMM = 41.7), and that iED were less confident that iDEP+ANX (EMM = 39.0; *p* < .05 in all cases). When including covariates, there were no significant effects (*F*s < 2.08, *ps* > .126).

##### Self-reported difficulty

In Bayesian analyses, the model including the transdiagnostic grouping was favored over the model that separated the patient groups (BF > 100). Grouping participants transdiagnostically without covariates, condition had a main effect on self-reported difficulty (*F*(2,566) = 14.39, *p* < .001), such that ratings were higher in the no-guessing condition (EMM = 65.3) than the other two HB conditions (guessing: EMM = 57.4, *t*(566) = 3.57; breath-hold: EMM = 53.6; *t*(566) = 5.25, *p* < .001 for both). When including covariates, there was only a marginal effect of condition (*F*(2,605) = 2.64, *p* = .072) such that the no-guessing condition was rated as more difficult than the other two conditions (*ps* < .001). When separating the groups by diagnosis, there was again an effect of condition (*F*(2,552) = 10.33, *p* < .001), with higher ratings in the no-guessing condition (EMM = 64.2; guessing: EMM = 56.9, *t*(552) = 3.53; breath-hold: EMM = 55.4; *t*(552) = 4.25, *p* < .001 in both cases), but no significant predictors were found after adding in covariates.

##### Heart rate

Bayes factor analyses favored the transdiagnostic model over one that separated patients by diagnosis (BF > 100). First, in an LME without covariates and grouping participants transdiagnostically, there was a main effect of condition (*F*(2,566) = 4.80, *p* = .008) such that heart rate was significantly higher in the breath-hold condition (EMM = 69.6) than the no-guessing condition (EMM = 66.6, *t*(566) = 3.09, *p* = .002). When including covariates (except number of heartbeats), there was again an effect of condition (*F*(2,566) = 4.80, *p* = .009) as well as a negative effect of PTT (*F*(1,277) = 5.93, *p* = .016). Post-hoc contrasts confirmed that heart rate was higher in the breath-hold condition than the no-guessing condition (*p* = .002). A similar pattern of results emerged when separating the patient groups by diagnosis. In a simple LME, there was an effect of condition (*F*(2,552) = 5.46, *p* = .005) with higher heart rates in the breath-hold condition (EMM = 69.5) than the no-guessing condition (EMM = 66.5, *t*(552) = 3.30, *p* = .001). After including covariates, condition (*F*(2,552) = 5.46, *p* = .005) and PTT (*F*(1,270) = 6.19, *p* = .013) predicted heart rate. Post-hoc contrasts revealed significantly greater heart rates in the breath-hold condition (EMM = 69.4) than the no-guessing condition (EMM = 66.4, *t*(552) = 3.30, *p* = .001) and PTT was again negatively associated with heart rate.

#### Associations with interoceptive awareness scales

Sensory precision (*IP*) in the no-guessing condition was positively associated with two MAIA subscales: Attention Regulation (*r* = .18, *p* = .01) and Trust (*r* = .13, *p* = .05). No relationships were found for *pHB* and any self-report measures; however, multiple relationships were found between *AvR* values and various interoceptive awareness scales. In the no-guessing condition, *AvR* values were negatively associated with Self-Regulation (*r* = -.15, *p* = .03). In the guessing condition, *AvR* values were negatively associated with MAIA Emotional Awareness (r = -.18, *p* = .01), Body-Listening (*r* = -.14, *p* = .04), and Not-Distracting (*r* = -.23, *p* < .001). Finally, *AvR* in the breath-hold condition was positively associated with MAIA Not-Worrying (*r* = .15, *p* = .02). However, none of these findings replicate the results from our prior study.

#### Sample comparisons and combined sample analyses

As reported in **Table S1** below, there was no evidence for differences in any measure between samples within each clinical group, with the exception of BMI, which was greater in iEDs in the confirmatory sample.

### Supplementary Tables

**Table S1.** Comparisons of characteristics between the exploratory and confirmatory samples (sample sizes given are the combined group sizes). Bayes Factors (BFs) are reported to compare the values between samples within each group.

| Individual difference variable* | Sample (and Bayes Factor comparison) | Healthy comparison (N = 111) | Anxiety (N = 19) | Depression (N = 95) | Depression + Anxiety (N = 241) | Eating Disorder (N = 38) | Substance use disorder (N = 215) |
| --- | --- | --- | --- | --- | --- | --- | --- |
| **Age** | Exploratory | 32.04 (11.08) | 36.42 (10.01) | 37.12 (11.83) | 35.28 (11.23) | 27.40 (9.83) | 34.03 (8.88) |
|  | Confirmatory | 33.29 (11.39) | 37.86 (3.85) | 36.66 (10.82) | 31.80 (10.48) | 28.06 (9.22) | 34.84 (8.52) |
|  | BF | 0.23 | 0.47 | 0.24 | 2.01 | 0.33 | 0.19 |
| **Sex** | Exploratory | 50% male | 33% male | 30% male | 26% male | 14% male | 45% male |
|  | Confirmatory | 42% male | 25% male | 42% male | 20% male | 8% male | 39% male |
|  | BF | 0.47 | 0.59 | 0.58 | 0.32 | 0.42 | 0.34 |
| **PHQ-9** | Exploratory | 0.83 (1.29) | 7.47 (5.80) | 13.48 (4.73) | 13.10 (5.21) | 12.93 (7.98) | 6.68 (5.91) |
|  | Confirmatory | 0.97 (1.76) | 7.50 (3.11) | 12.12 (5.13) | 12.95 (5.09) | 13.17 (6.65) | 6.50 (5.77) |
|  | BF | 0.22 | 0.46 | 0.45 | 0.15 | 0.33 | 0.16 |
| **OASIS** | Exploratory | 1.37 (1.89) | 10.60 (2.16) | 7.55 (3.34) | 10.51 (3.12) | 9.50 (4.93) | 5.94 (4.77) |
|  | Confirmatory | 1.02 (1.51) | 9.00 (1.41) | 7.19 (3.44) | 10.36 (3.14) | 10.04 (4.28) | 5.70 (4.10) |
|  | BF | 0.34 | 0.83 | 0.26 | 0.15 | 0.34 | 0.16 |
| **DAST-10** | Exploratory | 0.10 (0.30) | 0.33 (0.49) | 0.72 (1.66) | 0.58 (1.13) | 1.23 (2.62) | 7.55 (2.05) |
|  | Confirmatory | 0.24 (0.57) | 0.00 (0.00) | 0.58 (1.17) | 0.47 (1.06) | 1.25 (1.59) | 7.19 (2.56) |
|  | BF | 0.64 | 0.80 | 0.26 | 0.19 | 0.33 | 0.28 |
| **PTT±** | Exploratory | 0.20 (0.02) | 0.20 (0.01) | 0.19 (0.01) | 0.20 (0.02) | 0.20 (0.02) | 0.20 (0.02) |
|  | Confirmatory | 0.19 (0.02) | 0.17 (0.04) | 0.19 (0.020) | 0.20 (0.03) | 0.20 (0.02) | 0.20 (0.03) |
|  | BF | 0.40 | 1.83 | 0.25 | 0.22 | 0.39 | 0.24 |
| **BMI** | Exploratory | 27.59 (5.54) | 27.05 (6.02) | 28.51 (5.47) | 28.77 (5.49) | 22.25 (4.43) | 28.23 (4.56) |
|  | Confirmatory | 28.15 (5.88) | 31.85 (3.38) | 27.96 (5.93) | 27.46 (5.52) | 24.76 (4.07) | 28.93 (5.28) |
|  | BF | 0.22 | 1.15 | 0.25 | 0.62 | **3.87** | 0.23 |

\*PHQ-9 = Patient Health Questionnaire 9; OASIS = Overall Anxiety Sensitivity and Impairment Scale; DAST-10 = Drug Abuse Screening Test; PTT = median pulse transit time; BMI = Body Mass Index.

**Table S2.** Logistic regressions in iSUD predicting substance use disorders compared to HCs (N = 111) in the combined sample.

| Substance Use Disorders |  |  |  |  |  |  |
| --- | --- | --- | --- | --- | --- | --- |
| Disorder | Condition | Effect | Estimate | SE | *z* | *p* |
| Only Stimulant | Guessing (N = 43) | IP | 2.57 | 3.86 | 0.67 | .506 |
|  |  | pHB | 0.76 | 1.25 | 0.61 | .544 |
|  | No-Guessing (N = 42) | IP | 2.94 | 3.78 | 0.78 | .437 |
|  |  | pHB | 0.31 | 1.43 | 0.21 | .831 |
|  | Breath-hold (N = 41) | IP | -9.38 | 4.26 | -2.20 | .028 |
|  |  | pHB | 1.24 | 1.63 | 0.76 | .445 |
| Stimulant | Guessing (N = 174) | IP | 0.83 | 2.98 | 0.28 | .780 |
|  |  | pHB | -0.06 | 0.73 | -0.08 | .940 |
|  | No-Guessing (N = 173) | IP | 0.122 | 2.80 | 0.04 | .965 |
|  |  | pHB | 1.86 | 0.98 | 1.89 | .058 |
|  | Breath-hold (N = 171) | IP | -11.56 | 2.77 | -4.18 | < .001 |
|  |  | pHB | 2.43 | 1.12 | 2.17 | .030 |
| Cannabis | Guessing (N = 97) | IP | 0.82 | 3.31 | 0.25 | .805 |
|  |  | pHB | -0.26 | 0.86 | -0.30 | .767 |
|  | No-Guessing (N = 97) | IP | -3.06 | 3.49 | -0.88 | .381 |
|  |  | pHB | 2.56 | 1.06 | 2.41 | .016 |
|  | Breath-hold (N = 96) | IP | -10.59 | 3.16 | -3.35 | < .001 |
|  |  | pHB | 2.01 | 1.25 | 1.61 | .108 |
| Opioid | Guessing (N = 77) | IP | 1.47 | 3.46 | 0.42 | .672 |
|  |  | pHB | 0.14 | 0.98 | 0.15 | .884 |
|  | No-Guessing (N = 77) | IP | -0.01 | 3.20 | 0.00 | .998 |
|  |  | pHB | 2.35 | 1.17 | 2.01 | .044 |
|  | Breath-hold (N = 76) | IP | -10.21 | 3.26 | -3.13 | .002 |
|  |  | pHB | 2.96 | 1.23 | 2.41 | .016 |
| Alcohol | Guessing (N = 74) | IP | 2.90 | 3.28 | 0.88 | .377 |
|  |  | pHB | -1.36 | 0.99 | -1.37 | .171 |
|  | No-Guessing (N = 74) | IP | -1.52 | 3.41 | -0.45 | .656 |
|  |  | pHB | 2.05 | 1.20 | 1.72 | .086 |
|  | Breath-hold (N = 73) | IP | -11.89 | 3.65 | -3.26 | .001 |
|  |  | pHB | 2.70 | 1.35 | 2.00 | .046 |
| Sedative | Guessing (N = 55) | IP | 2.04 | 3.82 | 0.53 | .594 |
|  |  | pHB | -0.45 | 1.06 | -0.42 | .675 |
|  | No-Guessing (N = 55) | IP | -2.57 | 3.84 | -0.67 | .504 |
|  |  | pHB | 3.01 | 1.21 | 2.48 | .013 |
|  | Breath-hold (N = 53) | IP | -8.43 | 3.49 | -2.42 | .016 |
|  |  | pHB | 2.63 | 1.36 | 1.93 | .054 |
| Affective Disorders (within SUDs) |  |  |  |  |  |  |
| MDD | Guessing (N = 127) | IP | 3.10 | 3.01 | 1.03 | .302 |
|  |  | pHB | 0.26 | 0.87 | 0.29 | .769 |
|  | No-Guessing (N = 127) | IP | 1.14 | 2.83 | 0.40 | .686 |
|  |  | pHB | 1.51 | 1.05 | 1.44 | .149 |
|  | Breath-hold (N = 127) | IP | -9.91 | 2.82 | -3.52 | < .001 |
|  |  | pHB | 1.91 | 1.15 | 1.67 | .096 |
| **GAD** | Guessing<br>(N = 41) | IP | -0.84 | 4.29 | -0.20 | .845 |
|  |  | pHB | 0.97 | 1.18 | 0.82 | .411 |
|  | No-Guessing<br>(N = 40) | IP | -3.20 | 4.51 | -0.71 | .478 |
|  |  | **pHB** | **3.63** | **1.32** | **2.75** | **.006** |
|  | Breath-hold<br>(N = 40) | **IP** | **-10.30** | **4.23** | **-2.43** | **.015** |
|  |  | pHB | 2.65 | 1.53 | 1.73 | .083 |
| **PTSD** | Guessing<br>(N = 32) | IP | 0.95 | 4.75 | 0.20 | .841 |
|  |  | pHB | -1.37 | 1.42 | 0.97 | .334 |
|  | No-Guessing<br>(N = 32) | IP | 0.53 | 4.20 | 0.13 | .900 |
|  |  | pHB | 1.05 | 1.48 | 0.71 | .479 |
|  | Breath-hold<br>(N = 31) | IP | -6.18 | 3.85 | -1.61 | .109 |
|  |  | pHB | 2.91 | 1.63 | 1.79 | .074 |
| **Social Anxiety** | Guessing<br>(N = 26) | IP | -4.87 | 5.98 | -0.81 | .416 |
|  |  | pHB | 2.56 | 1.45 | 1.76 | .078 |
|  | No-Guessing<br>(N = 26) | IP | -5.62 | 5.76 | -0.98 | .329 |
|  |  | pHB | 1.65 | 1.48 | 1.11 | .267 |
|  | Breath-hold<br>(N = 26) | **IP** | **-16.96** | **6.59** | **-2.57** | **.010** |
|  |  | pHB | 2.32 | 1.91 | 1.22 | .224 |
| **Panic** | Guessing<br>(N = 21) | IP | -1.79 | 5.97 | -0.30 | .765 |
|  |  | pHB | 1.81 | 1.65 | 1.10 | .270 |
|  | No-Guessing<br>(N = 21) | IP | -6.85 | 6.70 | -1.02 | .306 |
|  |  | pHB | 0.88 | 1.68 | 0.53 | .599 |
|  | Breath-hold<br>(N = 22) | **IP** | **-13.20** | **6.10** | **-2.16** | **.030** |
|  |  | pHB | 2.03 | 1.90 | 1.07 | .284 |

**Note.** Sample sizes within diagnostic groups are not equal across conditions because some participants did not have valid data in all conditions. We did not test the group of iSUDs with hallucinogen use disorder against HCs due to small sample size (8 exploratory, 1 confirmatory).

**Table S3.**
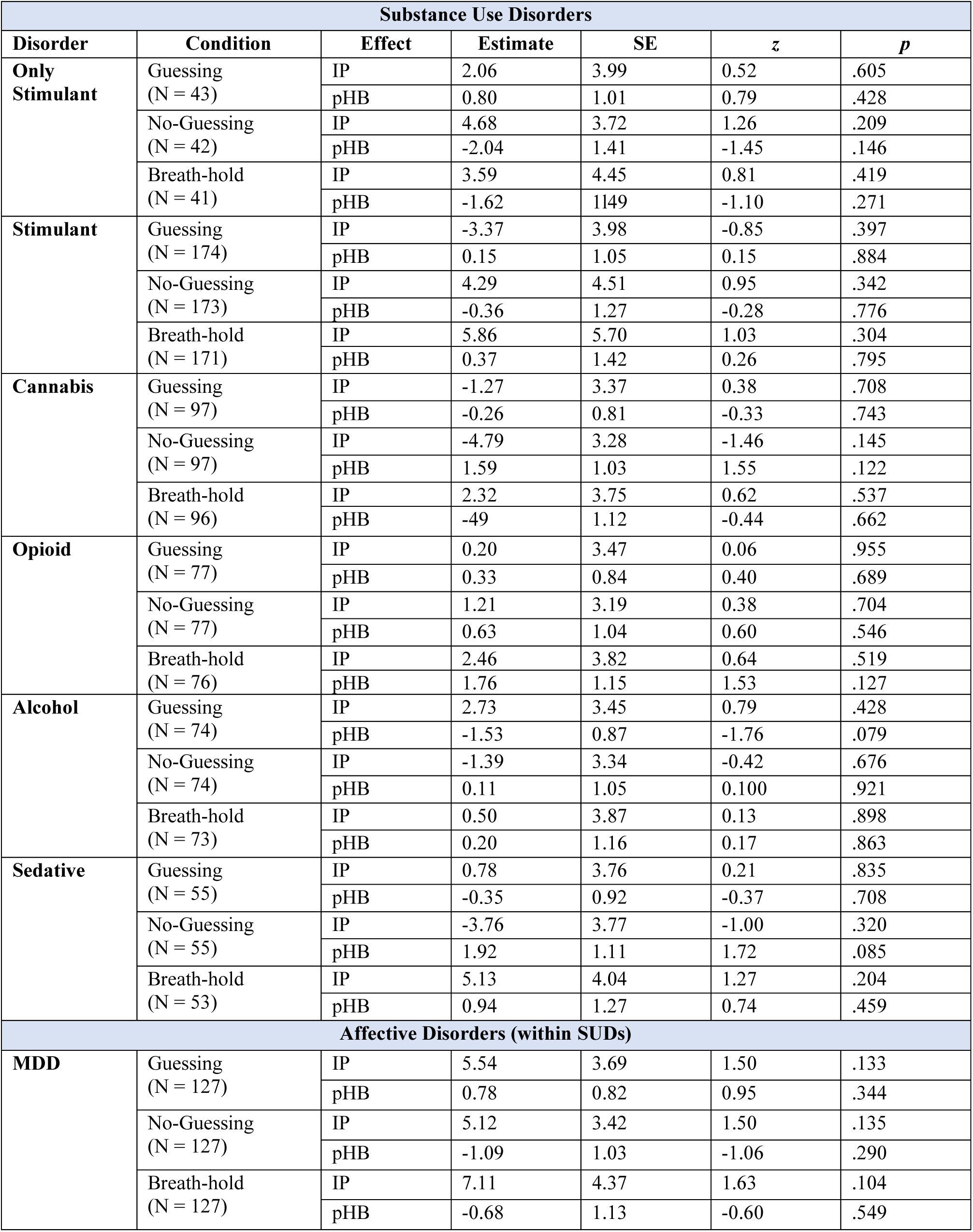
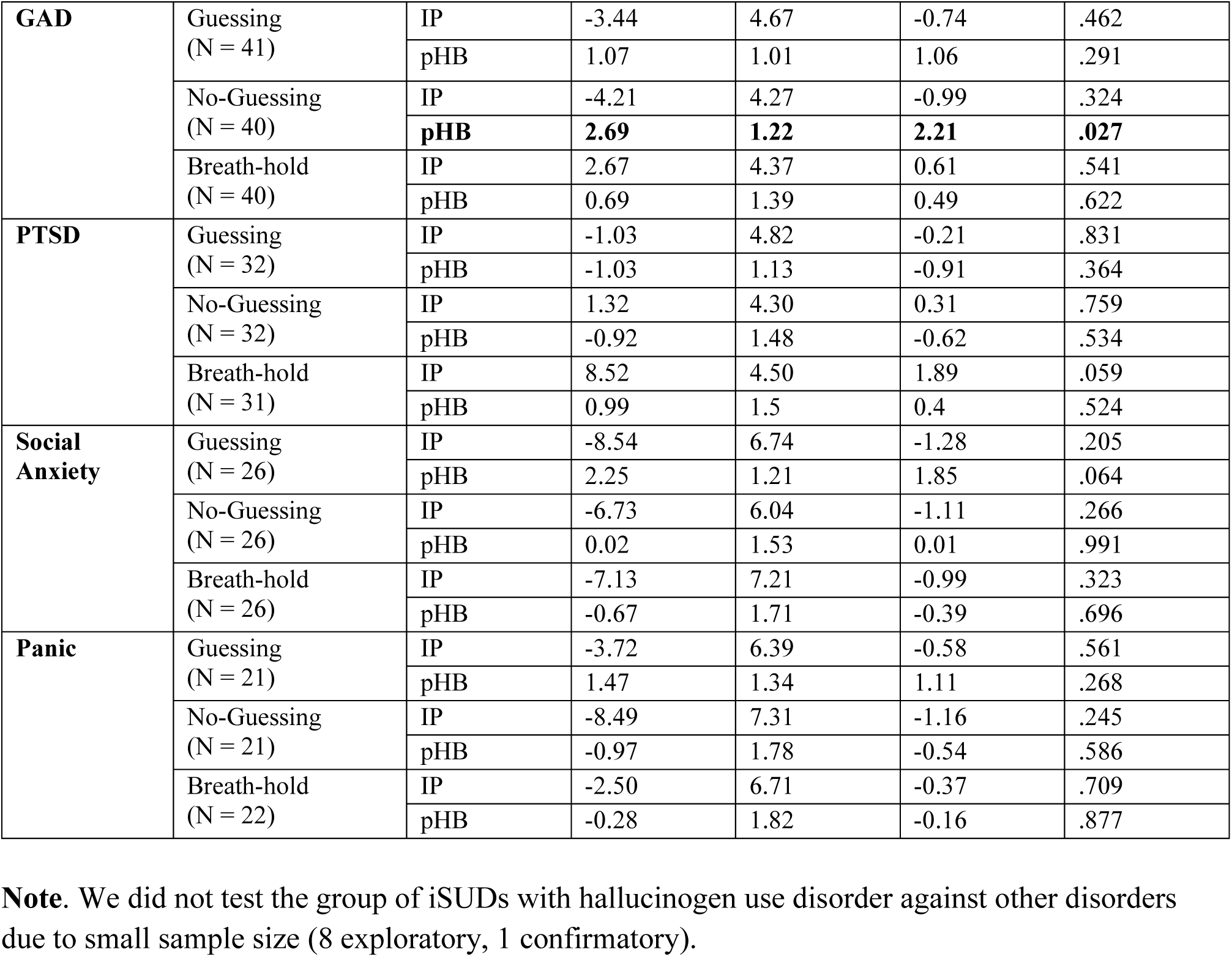
Logistic regressions in iSUD (N = 215) predicting each substance use disorder compared to all other disorders in the combined sample.

| Substance Use Disorders |  |  |  |  |  |  |
| --- | --- | --- | --- | --- | --- | --- |
| Disorder | Condition | Effect | Estimate | SE | *z* | *p* |
| Only Stimulant | Guessing (N = 43) | IP | 2.06 | 3.99 | 0.52 | .605 |
|  |  | pHB | 0.80 | 1.01 | 0.79 | .428 |
|  | No-Guessing (N = 42) | IP | 4.68 | 3.72 | 1.26 | .209 |
|  |  | pHB | -2.04 | 1.41 | -1.45 | .146 |
|  | Breath-hold (N = 41) | IP | 3.59 | 4.45 | 0.81 | .419 |
|  |  | pHB | -1.62 | 1.149 | -1.10 | .271 |
| Stimulant | Guessing (N = 174) | IP | -3.37 | 3.98 | -0.85 | .397 |
|  |  | pHB | 0.15 | 1.05 | 0.15 | .884 |
|  | No-Guessing (N = 173) | IP | 4.29 | 4.51 | 0.95 | .342 |
|  |  | pHB | -0.36 | 1.27 | -0.28 | .776 |
|  | Breath-hold (N = 171) | IP | 5.86 | 5.70 | 1.03 | .304 |
|  |  | pHB | 0.37 | 1.42 | 0.26 | .795 |
| Cannabis | Guessing (N = 97) | IP | -1.27 | 3.37 | 0.38 | .708 |
|  |  | pHB | -0.26 | 0.81 | -0.33 | .743 |
|  | No-Guessing (N = 97) | IP | -4.79 | 3.28 | -1.46 | .145 |
|  |  | pHB | 1.59 | 1.03 | 1.55 | .122 |
|  | Breath-hold (N = 96) | IP | 2.32 | 3.75 | 0.62 | .537 |
|  |  | pHB | -49 | 1.12 | -0.44 | .662 |
| Opioid | Guessing (N = 77) | IP | 0.20 | 3.47 | 0.06 | .955 |
|  |  | pHB | 0.33 | 0.84 | 0.40 | .689 |
|  | No-Guessing (N = 77) | IP | 1.21 | 3.19 | 0.38 | .704 |
|  |  | pHB | 0.63 | 1.04 | 0.60 | .546 |
|  | Breath-hold (N = 76) | IP | 2.46 | 3.82 | 0.64 | .519 |
|  |  | pHB | 1.76 | 1.15 | 1.53 | .127 |
| Alcohol | Guessing (N = 74) | IP | 2.73 | 3.45 | 0.79 | .428 |
|  |  | pHB | -1.53 | 0.87 | -1.76 | .079 |
|  | No-Guessing (N = 74) | IP | -1.39 | 3.34 | -0.42 | .676 |
|  |  | pHB | 0.11 | 1.05 | 0.100 | .921 |
|  | Breath-hold (N = 73) | IP | 0.50 | 3.87 | 0.13 | .898 |
|  |  | pHB | 0.20 | 1.16 | 0.17 | .863 |
| Sedative | Guessing (N = 55) | IP | 0.78 | 3.76 | 0.21 | .835 |
|  |  | pHB | -0.35 | 0.92 | -0.37 | .708 |
|  | No-Guessing (N = 55) | IP | -3.76 | 3.77 | -1.00 | .320 |
|  |  | pHB | 1.92 | 1.11 | 1.72 | .085 |
|  | Breath-hold (N = 53) | IP | 5.13 | 4.04 | 1.27 | .204 |
|  |  | pHB | 0.94 | 1.27 | 0.74 | .459 |
| Affective Disorders (within SUDs) |  |  |  |  |  |  |
| MDD | Guessing (N = 127) | IP | 5.54 | 3.69 | 1.50 | .133 |
|  |  | pHB | 0.78 | 0.82 | 0.95 | .344 |
|  | No-Guessing (N = 127) | IP | 5.12 | 3.42 | 1.50 | .135 |
|  |  | pHB | -1.09 | 1.03 | -1.06 | .290 |
|  | Breath-hold (N = 127) | IP | 7.11 | 4.37 | 1.63 | .104 |
|  |  | pHB | -0.68 | 1.13 | -0.60 | .549 |
| **GAD** | Guessing<br>(N = 41) | IP | -3.44 | 4.67 | -0.74 | .462 |
|  |  | pHB | 1.07 | 1.01 | 1.06 | .291 |
|  | No-Guessing<br>(N = 40) | IP | -4.21 | 4.27 | -0.99 | .324 |
|  |  | **pHB** | **2.69** | **1.22** | **2.21** | **.027** |
|  | Breath-hold<br>(N = 40) | IP | 2.67 | 4.37 | 0.61 | .541 |
|  |  | pHB | 0.69 | 1.39 | 0.49 | .622 |
| **PTSD** | Guessing<br>(N = 32) | IP | -1.03 | 4.82 | -0.21 | .831 |
|  |  | pHB | -1.03 | 1.13 | -0.91 | .364 |
|  | No-Guessing<br>(N = 32) | IP | 1.32 | 4.30 | 0.31 | .759 |
|  |  | pHB | -0.92 | 1.48 | -0.62 | .534 |
|  | Breath-hold<br>(N = 31) | IP | 8.52 | 4.50 | 1.89 | .059 |
|  |  | pHB | 0.99 | 1.5 | 0.4 | .524 |
| **Social Anxiety** | Guessing<br>(N = 26) | IP | -8.54 | 6.74 | -1.28 | .205 |
|  |  | pHB | 2.25 | 1.21 | 1.85 | .064 |
|  | No-Guessing<br>(N = 26) | IP | -6.73 | 6.04 | -1.11 | .266 |
|  |  | pHB | 0.02 | 1.53 | 0.01 | .991 |
|  | Breath-hold<br>(N = 26) | IP | -7.13 | 7.21 | -0.99 | .323 |
|  |  | pHB | -0.67 | 1.71 | -0.39 | .696 |
| **Panic** | Guessing<br>(N = 21) | IP | -3.72 | 6.39 | -0.58 | .561 |
|  |  | pHB | 1.47 | 1.34 | 1.11 | .268 |
|  | No-Guessing<br>(N = 21) | IP | -8.49 | 7.31 | -1.16 | .245 |
|  |  | pHB | -0.97 | 1.78 | -0.54 | .586 |
|  | Breath-hold<br>(N = 22) | IP | -2.50 | 6.71 | -0.37 | .709 |
|  |  | pHB | -0.28 | 1.82 | -0.16 | .877 |

**Note.** We did not test the group of iSUDs with hallucinogen use disorder against other disorders due to small sample size (8 exploratory, 1 confirmatory).

**Table S4.** Logistic regressions predicting affective disorders in the iDEP, iANX, and iDEP/ANX compared to HCs (N = 111) in the combined sample.

| Affective Disorders |  |  |  |  |  |  |
| --- | --- | --- | --- | --- | --- | --- |
| Disorder | Condition | Effect | Estimate | SE | z | p |
| **MDD** | Guessing<br>(N = 328) | IP | 1.48 | 2.74 | 0.54 | .589 |
|  |  | pHB | -0.84 | 0.67 | -1.26 | .207 |
|  | No-Guessing<br>(N = 328) | IP | -3.66 | 2.66 | -1.38 | .169 |
|  |  | pHB | 0.47 | 0.85 | 0.56 | .576 |
|  | Breath-hold<br>(N = 325) | **IP** | **-8.05** | **2.13** | **-3.78** | **< .001** |
|  |  | pHB | 0.06 | 0.94 | 0.06 | .952 |
| **GAD** | Guessing<br>(N = 165) | IP | 1.98 | 3.03 | 0.65 | .514 |
|  |  | pHB | -1.17 | 0.80 | -1.46 | .144 |
|  | No-Guessing<br>(N = 165) | IP | -0.57 | 2.87 | -0.20 | .842 |
|  |  | pHB | 0.03 | 0.94 | 0.03 | .974 |
|  | Breath-hold<br>(N = 164) | **IP** | **-7.69** | **2.44** | **-3.15** | **.002** |
|  |  | pHB | 0.32 | 1.05 | 0.30 | .761 |
| **Social Anxiety** | Guessing<br>(N = 82) | IP | 2.90 | 3.40 | 0.85 | .394 |
|  |  | pHB | -0.26 | 0.93 | -0.28 | .776 |
|  | No-Guessing<br>(N = 82) | IP | -3.85 | 3.83 | -1.01 | .315 |
|  |  | pHB | -0.92 | 1.18 | -0.79 | .430 |
|  | Breath-hold<br>(N = 81) | IP | -3.00 | 2.75 | -1.09 | .275 |
|  |  | pHB | -1.48 | 1.36 | -1.09 | .277 |
| **Panic** | Guessing<br>(N = 69) | IP | 0.98 | 3.55 | 0.28 | .783 |
|  |  | pHB | -0.42 | 0.95 | -0.44 | .657 |
|  | No-Guessing<br>(N = 69) | **IP** | **-9.14** | **4.53** | **-2.02** | **.044** |
|  |  | pHB | 1.98 | 1.18 | 1.67 | .094 |
|  | Breath-hold<br>(N = 68) | **IP** | **-7.68** | **3.22** | **-2.38** | **.017** |
|  |  | pHB | 0.60 | 1.31 | 0.46 | .646 |
| **PTSD** | Guessing<br>(N = 63) | IP | 1.13 | 3.52 | 0.32 | .749 |
|  |  | pHB | -1.35 | 0.98 | -1.37 | .170 |
|  | No-Guessing<br>(N = 63) | IP | -4.26 | 4.03 | -1.06 | .291 |
|  |  | pHB | 1.06 | 1.14 | 0.93 | .351 |
|  | Breath-hold<br>(N = 63) | **IP** | **-9.01** | **3.42** | **-2.63** | **.008** |
|  |  | pHB | 1.42 | 1.28 | 1.11 | .267 |

**Table S5.** Logistic regressions predicting affective disorders in the DEP, ANX, and DEP/ANX groups (N = 355) compared to all other disorders (combined sample).

| Affective Disorders |  |  |  |  |  |  |
| --- | --- | --- | --- | --- | --- | --- |
| Disorder | Condition | Effect | Estimate | SE | z | p |
| **MDD** | Guessing<br>(N = 328) | IP | 3.25 | 5.70 | 0.57 | .568 |
|  |  | pHB | 1.20 | 1.20 | 1.01 | .315 |
|  | No-Guessing<br>(N = 328) | IP | -0.19 | 5.72 | -0.03 | .973 |
|  |  | pHB | 1.92 | 1.78 | 1.08 | .282 |
|  | Breath-hold<br>(N = 325) | IP | 0.78 | 4.84 | 0.16 | .872 |
|  |  | pHB | 0.83 | 1.74 | 0.48 | .634 |
| **GAD** | Guessing<br>(N = 165) | IP | 1.21 | 2.74 | 0.44 | .659 |
|  |  | pHB | -0.16 | 0.63 | -0.25 | .805 |
|  | No-Guessing<br>(N = 165) | **IP** | **6.93** | **3.00** | **2.31** | **.021** |
|  |  | pHB | -0.58 | 0.82 | -0.72 | .474 |
|  | Breath-hold<br>(N = 164) | IP | -0.09 | 2.46 | -0.04 | .970 |
|  |  | pHB | 0.35 | 0.87 | 0.40 | .686 |
| **Social Anxiety** | Guessing<br>(N = 82) | IP | 2.61 | 3.11 | 0.84 | .402 |
|  |  | pHB | 0.86 | 0.75 | 1.15 | .250 |
|  | No-Guessing<br>(N = 82) | IP | -0.80 | 3.56 | -0.23 | .822 |
|  |  | pHB | -1.81 | 1.05 | -1.72 | .085 |
|  | Breath-hold<br>(N = 81) | **IP** | **6.01** | **2.75** | **2.18** | **.029** |
|  |  | pHB | -1.82 | 1.16 | -1.57 | .117 |
| **Panic** | Guessing<br>(N = 69) | IP | -0.27 | 3.46 | -0.08 | .939 |
|  |  | pHB | 0.62 | 0.79 | 0.78 | .435 |
|  | No-Guessing<br>(N = 69) | IP | -6.16 | 4.07 | -1.52 | .130 |
|  |  | pHB | 1.72 | 0.95 | 1.81 | .071 |
|  | Breath-hold<br>(N = 68) | IP | -0.02 | 3.09 | -0.01 | .996 |
|  |  | pHB | 0.46 | 1.07 | 0.43 | .671 |
| **PTSD** | Guessing<br>(N = 63) | IP | 0.49 | 3.55 | 0.14 | .891 |
|  |  | pHB | -0.45 | 0.83 | -0.55 | .585 |
|  | No-Guessing<br>(N = 63) | IP | -0.86 | 3.81 | -0.22 | .822 |
|  |  | pHB | 0.96 | 1.00 | 0.96 | .338 |
|  | Breath-hold<br>(N = 63) | IP | -1.39 | 3.26 | -0.43 | .670 |
|  |  | pHB | 1.36 | 1.06 | 1.28 | .199 |

**Table S6.** Logistic regressions predicting affective disorders in the eating disorders group compared to HCs (N = 111) in the combined sample.

| Eating Disorders |  |  |  |  |  |  |
| --- | --- | --- | --- | --- | --- | --- |
| Disorder | Condition | Effect | Estimate | SE | *z* | *p* |
| **Anorexia Nervosa** | Guessing (N = 10) | IP | 4.48 | 6.72 | 0.67 | .505 |
|  |  | pHB | -2.20 | 2.49 | -0.88 | .378 |
|  | No-Guessing (N = 10) | IP | 7.40 | 7.09 | 1.04 | .297 |
|  |  | pHB | -4.88 | 4.12 | -1.19 | .236 |
|  | Breath-hold (N = 10) | IP | -1.24 | 7.33 | -0.17 | .866 |
|  |  | pHB | -6.07 | 4.33 | -1.40 | .161 |
| **Bulimia / Binge Eating** | Guessing (N = 22) | IP | 6.30 | 4.21 | 1.50 | .135 |
|  |  | pHB | 1.34 | 1.65 | 0.81 | .418 |
|  | No-Guessing (N = 22) | IP | -3.39 | 5.84 | -0.58 | .562 |
|  |  | pHB | 0.51 | 1.83 | 0.28 | .780 |
|  | Breath-hold (N = 22) | IP | -8.15 | 5.13 | -1.59 | .112 |
|  |  | pHB | 0.77 | 1.98 | 0.39 | .697 |

**Table S7.** Logistic regressions predicting eating disorders in the Eating group (N = 38) compared to those without each disorder (combined sample).

| Eating Disorders |  |  |  |  |  |  |
| --- | --- | --- | --- | --- | --- | --- |
| Disorder | Condition | Effect | Estimate | SE | *z* | *p* |
| **Anorexia Nervosa** | Guessing (N = 10) | IP | 0.28 | 7.19 | 0.04 | .969 |
|  |  | pHB | -1.80 | 2.40 | -0.75 | .455 |
|  | No-Guessing (N = 10) | IP | 6.46 | 7.32 | 0.88 | .377 |
|  |  | pHB | -6.42 | 4.42 | -1.45 | .147 |
|  | Breath-hold (N = 10) | IP | 4.47 | 8.20 | 0.55 | .585 |
|  |  | pHB | -5.54 | 4.25 | -1.30 | .192 |
| **Bulimia / Binge Eating** | Guessing (N = 22) | IP | 6.61 | 7.56 | 0.88 | .382 |
|  |  | pHB | 3.29 | 2.26 | 1.46 | .145 |
|  | No-Guessing (N = 22) | IP | -10.50 | 7.31 | -1.44 | .151 |
|  |  | pHB | 2.29 | 3.21 | 0.71 | .476 |
|  | Breath-hold (N = 22) | IP | -1.94 | 7.60 | -0.26 | .798 |
|  |  | pHB | 3.91 | 3.19 | 1.22 | .221 |

## References

1. Khalsa SS, Adolphs R, Cameron OG, Critchley HD, Davenport PW, Feinstein JS, et al. Interoception and Mental Health: A Roadmap. Biol Psychiatry Cogn Neurosci Neuroimaging. 2018;3(6):501–13.

2. Furman DJ, Waugh CE, Bhattacharjee K, Thompson RJ, Gotlib IH. Interoceptive awareness, positive affect, and decision making in major depressive disorder. J Affect Disord. 2013;151(2):780–5.

3. Terhaar J, Viola FC, Bar KJ, Debener S. Heartbeat evoked potentials mirror altered body perception in depressed patients. Clin Neurophysiol. 2012;123(10):1950–7.

4. Dunn BD, Stefanovitch I, Evans D, Oliver C, Hawkins A, Dalgleish T. Can you feel the beat? Interoceptive awareness is an interactive function of anxiety-and depression-specific symptom dimensions. Behav Res Ther. 2010;48(11):1133–8.

5. Pollatos O, Traut-Mattausch E, Schandry R. Differential effects of anxiety and depression on interoceptive accuracy. Depress Anxiety. 2009;26(2):167–73.

6. Khalsa SS, Lapidus RC. Can Interoception Improve the Pragmatic Search for Biomarkers in Psychiatry? Front Psychiatry. 2016;7:121.

7. Stewart JL, Khalsa SS, Kuplicki R, Puhl M, Investigators T, Paulus MP. Interoceptive attention in opioid and stimulant use disorder. Addict Biol. 2020;25(6):e12831.

8. Khalsa SS, Craske MG, Li W, Vangala S, Strober M, Feusner JD. Altered interoceptive awareness in anorexia nervosa: Effects of meal anticipation, consumption and bodily arousal. Int J Eat Disord. 2015;48(7):889–97.

9. Desmedt O, Luminet O, Corneille O. The heartbeat counting task largely involves non-interoceptive processes: Evidence from both the original and an adapted counting task. Biol Psychol. 2018;138:185–8.

10. Phillips GC, Jones GE, Rieger EJ, Snell JB. Effects of the presentation of false heart-rate feedback on the performance of two common heartbeat-detection tasks. Psychophysiology. 1999;36(4):504–10.

11. Ring C, Brener J, Knapp K, Mailloux J. Effects of heartbeat feedback on beliefs about heart rate and heartbeat counting: a cautionary tale about interoceptive awareness. Biol Psychol. 2015;104:193–8.

12. Windmann S, Schonecke OW, Frohlig G, Maldener G. Dissociating beliefs about heart rates and actual heart rates in patients with cardiac pacemakers. Psychophysiology. 1999;36(3):339–42.

13. Ring C, Brener J. Heartbeat counting is unrelated to heartbeat detection: A comparison of methods to quantify interoception. Psychophysiology. 2018;55(9):e13084.

14. Murphy J, Millgate E, Geary H, Ichijo E, Coll MP, Brewer R, et al. Knowledge of resting heart rate mediates the relationship between intelligence and the heartbeat counting task. Biol Psychol. 2018;133:1–3.

15. Smith R, Kuplicki R, Feinstein J, Forthman KL, Stewart JL, Paulus MP, et al. A Bayesian computational model reveals a failure to adapt interoceptive precision estimates across depression, anxiety, eating, and substance use disorders. PLoS Computational Biology. 2020;16(12):e1008484.

16. Smith R, Kuplicki R, Teed A, Upshaw V, Khalsa SS. Confirmatory Evidence that Healthy Individuals Can Adaptively Adjust Prior Expectations and Interoceptive Precision Estimates. In: Verbelen T, Lanillos P, Buckley C, De Boom C, editors. Active Inference. Communications in Computer and Information Science. Communications in Computer and Information Science, vol 1326: Springer, Cham.; 2020. p. 156–64.

17. Khalsa SS, Rudrauf D, Sandesara C, Olshansky B, Tranel D. Bolus isoproterenol infusions provide a reliable method for assessing interoceptive awareness. Int J Psychophysiol. 2009;72(1):34–45.

18. Hassanpour MS, Yan L, Wang DJ, Lapidus RC, Arevian AC, Simmons WK, et al. How the heart speaks to the brain: neural activity during cardiorespiratory interoceptive stimulation. Philos Trans R Soc Lond B Biol Sci. 2016;371(1708).

19. Schandry R, Bestler M, Montoya P. On the relation between cardiodynamics and heartbeat perception. Psychophysiology. 1993;30(5):467–74.

20. Victor TA, Khalsa SS, Simmons WK, Feinstein JS, Savitz J, Aupperle RL, et al. Tulsa 1000: a naturalistic study protocol for multilevel assessment and outcome prediction in a large psychiatric sample. BMJ Open. 2018;8(1):e016620.

21. Kroenke K, Spitzer RL, Williams JB. The PHQ-9: validity of a brief depression severity measure. J Gen Intern Med. 2001;16(9):606–13.

22. Norman SB, Cissell SH, Means-Christensen AJ, Stein MB. Development and validation of an Overall Anxiety Severity And Impairment Scale (OASIS). Depress Anxiety. 2006;23(4):245–9.

23. Staley D, el-Guebaly N. Psychometric properties of the Drug Abuse Screening Test in a psychiatric patient population. Addict Behav. 1990;15(3):257–64.

24. Morgan JF, Reid F, Lacey JH. The SCOFF questionnaire: a new screening tool for eating disorders. West J Med. 2000;172(3):164–5.

25. Sheehan DV, Lecrubier Y, Sheehan KH, Amorim P, Janavs J, Weiller E, et al. The Mini-International Neuropsychiatric Interview (M.I.N.I.): the development and validation of a structured diagnostic psychiatric interview for DSM-IV and ICD-10. The Journal of clinical psychiatry. 1998;59 Suppl 20:22–33;quiz 4-57.

26. Ludwick-Rosenthal R, Neufeld RW. Heart beat interoception: a study of individual differences. Int J Psychophysiol. 1985;3(1):57–65.

27. Canales-Johnson A, Silva C, Huepe D, Rivera-Rei A, Noreika V, Garcia Mdel C, et al. Auditory Feedback Differentially Modulates Behavioral and Neural Markers of Objective and Subjective Performance When Tapping to Your Heartbeat. Cereb Cortex. 2015;25(11):4490–503.

28. Murphy J, Brewer R, Coll M-P, Plans D, Hall M, Shiu SS, et al. I feel it in my finger: Measurement device affects cardiac interoceptive accuracy. Biological psychology. 2019;148:107765.

29. Allen J, Murray A. Age-related changes in the characteristics of the photoplethysmographic pulse shape at various body sites. Physiol Meas. 2003;24(2):297–307.

30. Friston K, Mattout J, Trujillo-Barreto N, Ashburner J, Penny W. Variational free energy and the Laplace approximation. Neuroimage. 2007;34(1):220–34.

31. R Core Team. R: A language and environment for statistical computing R Foundation for Statistical Computing. Vienna, Austria. 2022.

32. Bates D, Maechler, M., Bolker, B., & Walker, S. Fitting Linear Mixed-Effects Models Using lme4. Journal of Statistical Software. 2015;67(1):1–48.

33. Schandry R. Heart beat perception and emotional experience. Psychophysiology. 1981;18(4):483–8.

34. Mehling WE, Price C, Daubenmier JJ, Acree M, Bartmess E, Stewart A. The Multidimensional Assessment of Interoceptive Awareness (MAIA). PLoS One. 2012;7(11):e48230.

35. Bagby RM, Taylor GJ, Parker JD. The Twenty-item Toronto Alexithymia Scale--II. Convergent, discriminant, and concurrent validity. J Psychosom Res. 1994;38(1):33–40.

36. Sandin B, Chorot P, McNally RJ. Anxiety sensitivity index: normative data and its differentiation from trait anxiety. Behav Res Ther. 2001;39(2):213–9.

37. Rigoux L, Stephan KE, Friston KJ, Daunizeau J. Bayesian model selection for group studies - revisited. Neuroimage. 2014;84:971–85.

38. Stephan KE, Penny WD, Daunizeau J, Moran RJ, Friston KJ. Bayesian model selection for group studies. Neuroimage. 2009;46(4):1004–17.

39. Parker JD, Taylor GJ, Bagby RM. The 20-Item Toronto Alexithymia Scale. III. Reliability and factorial validity in a community population. J Psychosom Res. 2003;55(3):269–75.

40. Hosmer Jr DW, Lemeshow S, Sturdivant RX. Applied logistic regression: John Wiley & Sons; 2013.

41. Zamariola G, Maurage P, Luminet O, Corneille O. Interoceptive accuracy scores from the heartbeat counting task are problematic: Evidence from simple bivariate correlations. Biol Psychol. 2018;137:12–7.

42. Gu X, Filbey F. A bayesian observer model of drug craving. JAMA psychiatry. 2017;74(4):419–20.

43. Kulkarni KR, O’Brien M, Gu X. Longing to Act: Bayesian Inference as a Framework for Craving in Behavioral Addiction. Addictive Behaviors. 2023:107752.

44. Critchley HD, Sherrill SP, Ewing DL, van Praag CG, Habash-Bailey H, Quadt L, et al. Cardiac interoception in patients accessing secondary mental health services: A transdiagnostic study. Autonomic Neuroscience. 2023;245:103072.

45. Teed AR, Feinstein JS, Puhl M, Lapidus RC, Upshaw V, Kuplicki RT, et al. Association of generalized anxiety disorder with autonomic hypersensitivity and blunted ventromedial prefrontal cortex activity during peripheral adrenergic stimulation: a randomized clinical trial. JAMA psychiatry. 2022;79(4):323–32.

46. Verdonk C, Teed AR, White EJ, Ren X, Stewart JL, Paulus MP, et al. Heartbeat-evoked neural response abnormalities in generalized anxiety disorder during peripheral adrenergic stimulation. medRxiv. 2023:2023.06. 09.23291166.

47. De la Cruz F, Teed AR, Lapidus RC, Upshaw V, Schumann A, Paulus MP, et al. Central autonomic network alterations in anorexia nervosa following peripheral adrenergic stimulation. Biological Psychiatry: Cognitive Neuroscience and Neuroimaging. 2023;8(7):720–30.

48. Lapidus RC, Puhl M, Kuplicki R, Stewart JL, Paulus MP, Rhudy JL, et al. Heightened affective response to perturbation of respiratory but not pain signals in eating, mood, and anxiety disorders. PLoS One. 2020;15(7):e0235346.

49. Berner LA, Simmons AN, Wierenga CE, Bischoff-Grethe A, Paulus MP, Bailer UF, et al. Altered anticipation and processing of aversive interoceptive experience among women remitted from bulimia nervosa. Neuropsychopharmacology. 2019;44(7):1265–73.

50. Flux M, Fine TH, Poplin T, Al Zoubi O, Schoenhals WA, Schettler J, et al. Exploring the acute cardiovascular effects of Floatation-REST. Frontiers in Neuroscience. 2022;16:995594.

51. Choquette EM, Flux MC, Moseman SE, Chappelle S, Naegele J, Upshaw V, et al. The Impact of Floatation Therapy on Body Image and Anxiety in Anorexia Nervosa: A Randomized Clinical Efficacy Trial. Available at SSRN 4285497.

52. Quadt L, Garfinkel SN, Mulcahy JS, Larsson DE, Silva M, Jones A-M, et al. Interoceptive training to target anxiety in autistic adults (ADIE): A single-center, superiority randomized controlled trial. EClinicalMedicine. 2021;39.

53. Opdensteinen KD, Schaan L, Pohl A, Schulz A, Domes G, Hechler T. Interoception in presch olers: New insights into its assessment and relations to emotion regulation and stress. Biological Psychology. 2021;165:108166.

54. Khalsa SS, Hassanpour MS, Strober M, Craske MG, Arevian AC, Feusner JD. Interoceptive anxiety and body representation in anorexia nervosa. Frontiers in psychiatry. 2018;9:444.

55. Mathys CD, Lomakina EI, Daunizeau J, Iglesias S, Brodersen KH, Friston KJ, et al. Uncertainty in perception and the Hierarchical Gaussian Filter. Frontiers in human neuroscience. 2014;8:825.

